# Cell-free chromatin epigenomic profiling enables non-invasive pancreatic cancer cell-state identification

**DOI:** 10.64898/2026.04.02.26349987

**Authors:** Karl Semaan, Marc Eid, Damien Vasseur, Gunsagar S. Gulati, Cibelle Lima, Elio Ibrahim, Ji-Heui Seo, John J. Canniff, Hunter Savignano, Alexander Jordan, Leigh Culnane, Noa Phillips, Rashad Nawfal, Aislyn Schalck, Andressa Dias Costa, Elizabeth A. Andrews, Emma C. Coleman, Talal El Zarif, Garyoung Gary Lee, Razane El Hajj Chehade, Ze Zhang, Gaelle Nafeh, Wassim Daoud Khatoun, Brady James, Zhenjie Jin, Paulo Da Silva Cordeiro, Brad Fortunato, David Peng, Christopher Vellano, Tim Heffernan, Antoine Hollebecque, Antoine Italiano, Brandon M. Huffman, James M. Cleary, Jacob E. Berchuck, Toni K. Choueiri, Kimberly J. Perez, Jonathan Nowak, Andrew J. Aguirre, Brian M. Wolpin, Sylvan C. Baca, Matthew L. Freedman, Harshabad Singh

**Author notes:** **Correspondence:** Harshabad Singh, Matthew L. Freedman, Sylvan C. Baca. Contributed equally as co-first authors (Karl Semaan, Marc Eid). Contributed equally as co-senior authors (Andrew J. Aguirre, Brian M. Wolpin, Sylvan C. Baca, Matthew L. Freedman, Harshabad Singh).

## Abstract

Classical and basal-like transcriptional subtypes of pancreatic ductal adenocarcinoma (PDAC) are prognostic and may predict response to different chemotherapy regimens and RAS inhibitors. Current subtyping methods rely on tissue biopsies and remain challenging to integrate into clinical workflows. Herein, we present a novel approach for non-invasive subtyping of PDAC based on epigenomic profiling of circulating tumor DNA (ctDNA). In a multi-omics cohort of patient-derived xenografts, we identify highly recurrent regulatory elements associated with classical and basal-like PDAC. We then demonstrate that these epigenomic signatures can identify PDAC subtype from plasma epigenomic profiling in a multi-institutional cohort of patients with metastatic PDAC and integrate information from circulating histone modifications and DNA methylation to develop the Pancreatic Integrated Epigenomic Score (PIES). PIES is concordant with tissue-based labels and captures transcriptional subtype heterogeneity observed within biopsies. Furthermore, it improves prognostication over tissue-based subtyping suggestive of the recovery of ground truth tumor biology from plasma ctDNA. Our work provides a proof-of-concept for a circulating biomarker that enables transcriptional subtyping and informs therapeutic decisions in pancreatic cancer.

**Significance:** Transcriptional subtyping of pancreatic cancer can improve prognostication and inform treatment selection. Current subtyping approaches rely on tissue biopsy, are challenging to implement in clinical practice, and are limited by tumor heterogeneity and sampling error. Herein, we introduce a cell-free DNA-based epigenomic assay capable of inferring pancreatic cancer subtypes noninvasively.

## Introduction

Pancreatic ductal adenocarcinoma (PDAC) is currently the third leading cause of cancer-related deaths in the United States, with a rising incidence (1,2). The prognosis of patients with PDAC remains poor, with a median survival of less than one year (1,3). This is largely due to the majority of diagnoses occurring at an advanced stage, development of early therapeutic resistance, and the lack of reliable biomarkers to inform treatment decisions (1,3–5).

Over the last decade, several studies have established that PDAC can be stratified into transcriptional subtypes (6–10). The initial classification by Collison et al. in 2011 introduced three transcriptional subtypes (classical, quasi-mesenchymal, exocrine-like), which has since evolved into a more widely adopted two-subtype framework: classical and basal-like (6). The basal-like subtype is characterized by poor differentiation, epithelial-to-mesenchymal transition (EMT), increased metastatic potential, and resistance to PDAC conventional chemotherapy treatments such as FOLFIRINOX (10–12). In a real-world cohort analysis of over 7,000 patients with PDAC, we showed that the basal-like transcriptional subtype is the most significant adverse prognostic biomarker in this disease (8). In addition, recent data suggest that these transcriptional subtypes also predict response, with the more aggressive basal-like tumors responding favorably to gemcitabine over 5FU-based chemotherapy in two different clinical trials (13–16) and to KRAS inhibitors in pre-clinical models (17,18) compared to classical tumors. Hence, clinical implementation of PDAC transcriptional subtyping could improve prognostication and treatment selection, leading to improved outcomes for patients.

Current PDAC subtyping methods rely on RNA or protein expression of canonical marker genes on tissue biopsy (5,14,16), yet none are yet fully integrated in clinical workflows. These limitations arise in part from logistical challenges surrounding tissue biopsy, especially in tenuous or frail patients, as well as technical constraints such as limited tissue yield and low tumor cellularity (19,20). Furthermore, a single tumor biopsy may fail to capture the heterogeneity intrinsic to PDAC, where tumors may exhibit mixed classical and basal-like components (5,21). Liquid biopsy using circulating tumor DNA (ctDNA) analysis is well-suited to address these challenges as it is minimally invasive, requires limited sample input, and has the ability to reflect the aggregate burden of the disease (22–25). Specifically, epigenomic profiling of circulating chromatin shed from the tumor can capture varied tumoral epigenomic states allowing blood-based cancer subtyping (26–29) and easier clinical adoption and longitudinal sampling over the course of a patient’s treatment (22–25,30). Herein, we develop **P**ancreatic **I**ntegrated **E**pigenomic **S**core (PIES), a novel ctDNA epigenomic assay for plasma-based PDAC classical:basal-like determination.

## Results

### Classical and basal-like PDAC subtypes exhibit distinct epigenomic profiles

We first sought to comprehensively characterize the epigenomic landscape of classical and basal-like PDAC. We performed this analysis on 28 PDAC PDXs that we obtained from two institutional biobanks. PDXs were selected over primary pancreatic tumors because primary tumors have a high stromal content making epithelial tumor characterization challenging. Moreover, larger tumor volumes can be obtained from PDX models. To determine the PDX transcriptional subtypes, we performed bulk RNA-sequencing and employed PurIST, which is a single-sample classifier for tumor subtyping in pancreatic cancer, and categorizes tumors along a spectrum of strong, likely, and lean (9). We classified 18 strong classical and 10 strong basal-like tumors, and the subtype was further confirmed using the Moffitt gene signature (7) **(Figure S1A)**. To develop subtype-specific epigenomic signatures, we performed chromatin immunoprecipitation and sequencing (ChIP-seq) for two histone post-translational modifications: H3K27ac and H3K4me3. H3K27ac is enriched at active gene promoters and enhancers (31), and H3K4me3 is enriched at active or bivalent gene promoters (32). We also performed assay of transposase accessible chromatin (ATAC-seq) to profile chromatin accessibility (33,34) and methylated CpG dinucleotide immunoprecipitation and sequencing (MeDIP-seq) for DNA methylation (35,36) **(Figure 1A-B; Table S1)**. As expected, classical PDAC tumors exhibited a gain of signals at promoters of canonical classical subtype marker genes, including *CLDN18*, *GATA6*, and *HNF1A* (5,12,37,38) **(Figure 1C)**. In contrast, basal tumors demonstrated increased signal in key regulators of EMT and basal-like genes including *KRT6A*, *PTGES*, and *SNAI2* (5,7,9,39,40) **(Figure 1C)**.

**Figure 1.**
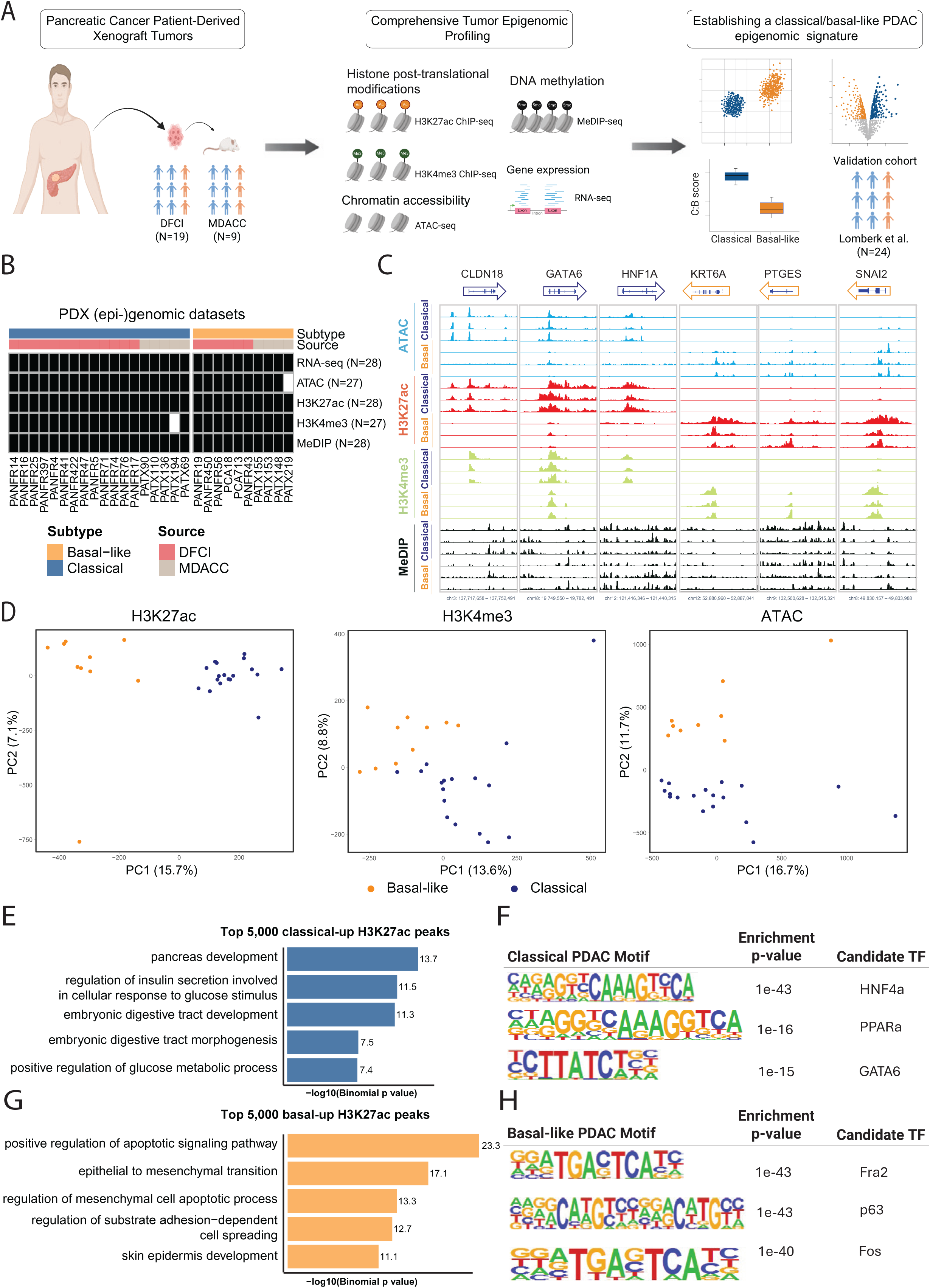
Comprehensive epigenomic profiling of classical and basal-like PDAC reveals widespread epigenomic differences. **(A)** Experimental framework for the epigenomic profiling of PDAC PDX. **(B)** Epigenomic datasets generated from PDAC PDX. **(C)** Representative epigenomic data from 3 classical and 3 basal-like PDAC PDX at classical associated (*CLDN18*, *GATA6*, *HNF1a*) and basal-like associated (*KRT6A*, *PTGES*, *SNAI2*) loci. **(D)** Principal component analysis (PCA) plots of the H3K27ac, H3K4me3, and ATAC peaks in classical and basal-like PDAC PDX. **(E)** GREAT analysis of classical-up H3K27ac peaks. **(F)** Significantly enriched nucleotide motifs present in classical-up sites by *de novo* motif analysis (all q-value<10^−5^). **(G)** GREAT analysis of basal-like-up H3K27ac peaks. **(H)** Significantly enriched nucleotide motifs present in basal-like-up sites by *de novo* motif analysis (all q-value<10^−5^).

To ascertain whether classical and basal-like tumors had subtype-specific epigenomic landscapes more globally, we conducted principal component analysis (PCA) and unsupervised hierarchical clustering (HC), for each epigenomic mark. Classical and basal-like PDAC models clearly separated when analyzing activated regions marked by H3K27ac, H3K4me3 and ATAC-seq peaks, whereas global DNA methylation was less discriminatory **(Figure 1D; Figure S1B-C)**.

We then sought to investigate the differential regions associated with classical and basal-like subtypes for each epigenomic feature (41). Using DESeq2, we identified 54,922 differentially marked H3K27ac regions (22,136 enriched in classical PDAC; “classical-up”; FDR-q<0.05; **Figure S1D; Table S2**), 8,754 differential H3K4me3 regions (3,808 classical-up; FDR-q<0.05), 48,088 differentially accessible ATAC-seq regions (21,479 classical-up peaks; FDR-q<0.05) and 752 differentially methylated regions on MeDIP-seq (442 classical-up; FDR-q<0.05). The H3K27ac classical-up sites were enriched for regions related to pancreas embryonic development and function **(Figure 1E)** and with motifs for transcription factors important for pancreatic lineage determination and known relevance to classical subtype biology, such as *GATA6* and *HNF4A* (39,42–44) **(Figure 1F)**. Conversely, H3K27ac sites upregulated in the basal-like subtype were enriched for pathways associated with epithelial-mesenchymal transition (EMT) and epidermal development **(Figure 1G)**, and p63 binding motifs **(Figure 1H)** consistent with earlier reports (8,37,45–47).

### Development of tumor-informed epigenomic signatures for classical and basal-like PDAC subtypes

We investigated the ability of these differential epigenomic regions to distinguish classical and basal-like PDAC subtypes in an independent set of previously published PDAC PDX models (39). We reanalyzed RNA-seq and ChIP-seq data from 24 external PDAC PDXs, identifying 17 strong classical PDAC and 3 strong basal-like PDXs, with the remainder categorized as “likely” or “lean” classical using PurIST (9) **(Table S3)**. Unsupervised hierarchical clustering **(Figure 2A)** and PCA **(Figure S2A)** of combined internal and previously published PDAC PDXs using H3K27ac signal clearly segregated the models by transcriptional subtype, despite technical differences in protocols and reagents utilized between our cohort and the external cohort. Furthermore, in the external validation PDX cohort, clear on-off patterns were observed across the differentially marked and accessible sites identified using our in-house PDX dataset **(Figure 2B)**. These results indicate highly recurrent epigenomic differences between PDAC transcriptional subtypes.

**Figure 2.**
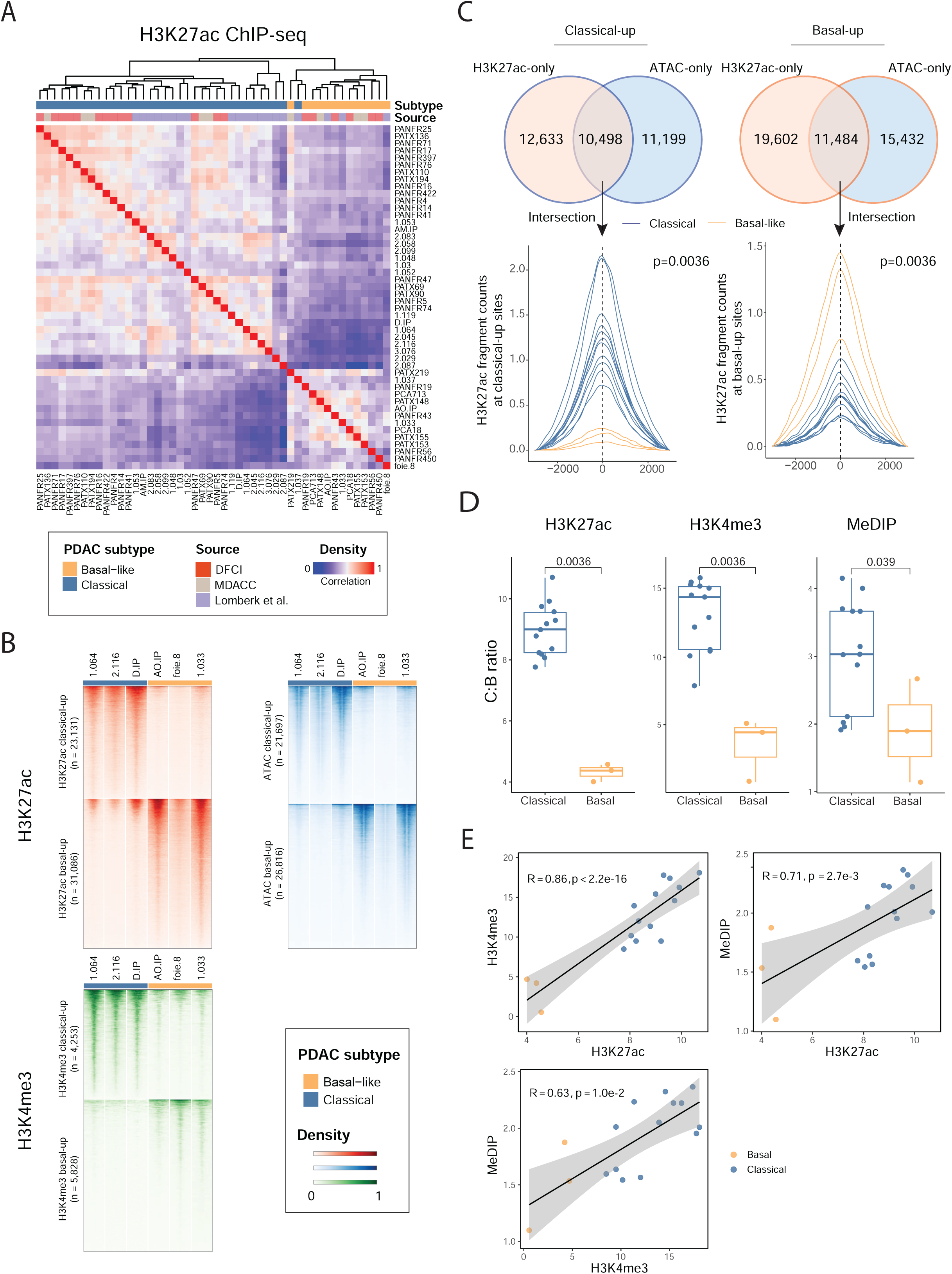
Developing epigenomic signatures for classical and basal-like PDAC subtypes. **(A)** Unsupervised hierarchical clustering of the H3K27ac peaks in a large set of PDX including an independent cohort of PDAC PDX. **(B)** Heatmap of normalized H3K27ac, ATAC, and H3K4me3 peaks tag densities (located ±2 kb from peak center) at subtype-specific differential sites in an independent cohort of PDAC PDX. **(C)** Framework for refining the epigenomic signature and evaluating its discriminatory power (Wilcoxon rank-sum test). **(D)** Boxplots showing signal at classical: basal-like (C:B) sites in an independent cohort of PDX (Wilcoxon rank-sum test). **(E)** Scatter plots of C:B signal ratios derived from H3K4me3 ChIP-seq, H3K27ac ChIP-seq, and MeDIP-seq in an independent cohort of PDAC PDX. Spearman correlation coefficients and corresponding p-values are shown.

We next sought to refine these differential epigenomic regions and reduce the number of sites while maintaining discrimination between PDAC subtypes. We first explored enhancer-centric signature by evaluating the intersection of H3K27ac and ATAC-seq differential regions, with the aim of capturing biologically relevant enhancers with higher accuracy (48). We categorized sites that were both differentially marked by H3K27ac and differentially accessible as “intersection” and compared their power of separation to that of unselected H3K27ac, unselected ATAC regions, “ATAC-only” regions (differentially accessible regions which are not differentially marked with H3K27ac), and “H3K27ac-only” (differential H3K27ac regions which are not differentially accessible; **Figure 2C; Methods**). For each site set, we quantified subtype separation as the absolute difference in per-site normalized aggregate signal between all pairwise combinations of classical and basal-like samples in the external validation cohort. “Intersection” sites (10,498 classical-up and 11,484 basal-up) demonstrated significantly higher separation of H3K27ac signal intensity between classical and basal-like PDAC subtypes compared to unselected H3K27ac (22,136 classical-up and 32,786 basal-up), unselected ATAC (21,479 classical-up and 26,609 basal-up), ATAC-only (11,199 classical-up and 15,432 basal-up) and H3K27ac-only sites (12,633 classical-up and 19,602 basal-up), for both classical and basal sites (all p<10^−12^; **Figure S2B-C**). Regarding the promoter-centric signature, increasing the stringency for the differential H3K4me3 ChIP-seq sites (FDR-q < 0.01 and log_2_ fold-change (LFC) > 2), retained 2,737 highly specific sites (977 classical-up and 1,760 basal-up sites), which led to the highest signal discrimination between the two subtypes compared to other LFC thresholds (all p<10^−10^; **Figure S2D-F**). For differentially methylated regions (DMRs) identified via MeDIP-seq, a more modest threshold of FDR-q < 0.01 and LFC > 1 was chosen given fewer differential regions overall, thereby retaining 274 subtype-specific DMRs (142 basal-like and 132 classical DMRs). Of note, the evaluation of MeDIP performance in the external PDX cohort may have been limited by data availability. Unlike the ChIP-seq datasets, raw DNA methylation sequencing data were not publicly accessible, which prevented us from performing read alignment and extracting fragment-level signals. Instead, methylation information was obtained as summarized scores at predefined genomic loci. As a result, we had to estimate the methylation signal by overlapping these loci with our subtype-specific DMRs, which led to the loss of a portion of sites.

To internally normalize the signature score for each individual mark, we calculated the signal ratio at classical:basal (C:B) sites. All three C:B epigenomic signature scores (H3K27ac, H3K4me3, MeDIP) were significantly different between classical and basal-like PDXs **(Figure 2D)** and were significantly correlated with each other in the external validation cohort **(Figure 2E)**. Hence, tumor-informed epigenomic signatures derived from our discovery institutional PDAC PDX cohort can broadly capture and distinguish classical and basal-like PDAC subtypes.

### PDX-informed epigenomic signatures enable plasma-based PDAC transcriptional subtype analysis

Next, we aimed to assess whether these tumor-informed epigenomic signatures could be recovered from plasma of patients with PDAC to discern their transcriptional subtype **(Figure 3A)**. We profiled cell-free DNA utilizing cell-free ChIP-seq (cfChIP-seq) for H3K27ac and H3K4me3 and cfMeDIP-seq similar to prior studies (25,49,23) on plasma of 82 patients with metastatic PDAC where tissue-based transcriptional subtype calls were available. The samples included patients from three distinct cohorts: A clinical trial of paricalcitol in combination with chemotherapy (NCT03520790, n=29), the MOSCATO-02 trial (NCT01566019, n=27), and an institutional cohort of patients treated with standard of care at the Dana-Farber Cancer Institute (n=26) **(Figure 3B; Table S4)**. The median age was 65 (IQR: 57—70), 58% were male, 97% had stage IV PDAC (two patients had stage III), and 68% were previously untreated. Our cohort included 50 classical and 32 basal-like samples based on RNA analysis of temporally matched tumor biopsy to the plasma collection. The median interval between the plasma draws and the tissue biopsy was 7 days (IQR: 1—15), with 97% of the biopsies obtained from a metastatic site, most commonly the liver **(Table S4)**. We performed cfChIP-seq for H3K27ac and H3K4me3 and cfMeDIP-seq on each patient’s plasma and generated a total of 242 cell-free epigenomic libraries. Circulating tumor DNA (ctDNA) fraction was estimated by applying ichorCNA (50). The median (IQR) ctDNA tumor fraction across these samples was 3.3% (0—9%), consistent with previous studies **(Figure S3A)**, and was comparable between classical and basal-like PDAC (3.2% vs. 4.8%, respectively; p=0.3) (51–53) **(Figure 3C)**.

**Figure 3.**
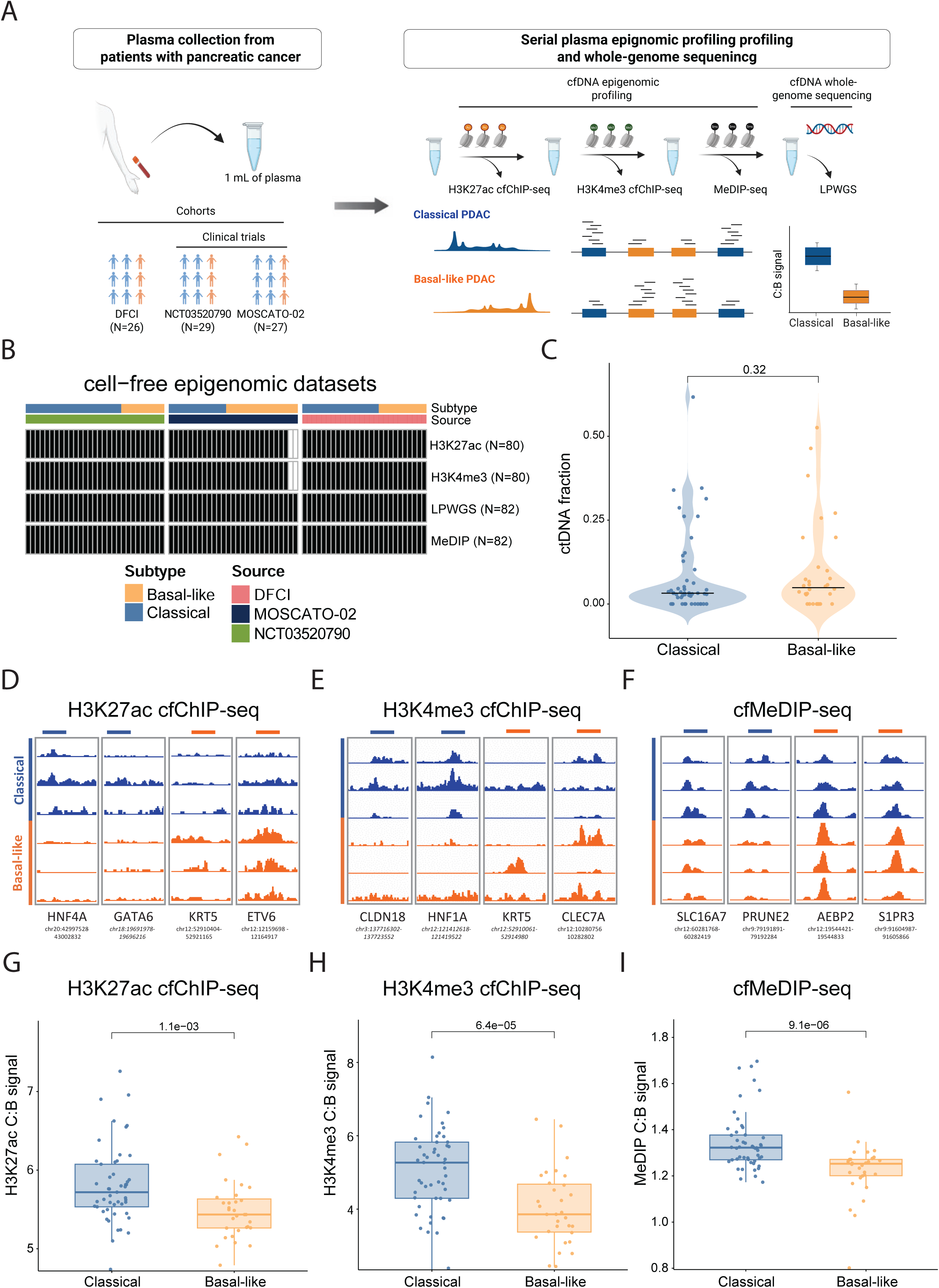
Cell-free DNA epigenomic profiling enables non-invasive subtyping of pancreatic cancer. **(A)** Overview of the experimental approach to perform multianalyte epigenomic profiling of cell-free DNA from 1 mL of patient plasma. **(B)** Cell-free epigenomic datasets generated from plasma of patients with metastatic pancreatic cancer. **(C)** ctDNA fraction distribution in plasma samples from patients’ pancreatic cancer by PDAC subtype (Wilcoxon rank-sum test). **(D)** H3K27ac cell-free (cf) ChIP profiles at tissue-informed differential sites between classical and basal-like subtypes. **(E)** H3K4me3 cfChIP profiles at tissue-informed differential sites between classical and basal-like subtypes. **(F)** cfMeDIP profiles at tissue-informed differential sites between classical and basal-like subtypes. **(G)** Boxplot showing H3K27ac cfChIP C:B ratio of aggregate signal at tissue-informed classical and basal-like specific enhancer-centric sites in plasma samples by PDAC subtype (Wilcoxon rank-sum test). **(H)** Boxplot showing H3K4me3 cfChIP aggregate signal at tissue-informed classical and basal-like specific promoter centric sites in plasma samples by PDAC subtype (Wilcoxon rank-sum test). **(I)** Boxplot showing cfMeDIP C:B ratio aggregate signal at tissue-informed classical and basal-like DMRs in plasma samples by PDAC subtype (Wilcoxon rank-sum test).

Notably, plasma H3K27ac, H3K4me3, and MeDIP signals were elevated at classical-up sites in classical PDAC samples and elevated at basal-like-up sites in basal-like PDAC samples **(Figure 3D-F)**. Similar to the analysis conducted on the PDX data, we calculated the normalized eigenomic signature scores (C:B ratios) at predefined signature sites from our PDX analysis for each epigenomic mark. C:B ratios were consistently higher in classical plasma samples compared to basal-like plasma samples for H3K27ac (p=1.1×10^−3^; **Figure 3G**), H3K4me3 (p=6.4×10^−5^; **Figure 3H**), and MeDIP (p=9.1×10^−6^; **Figure 3I**). Notably, C:B signal ratios derived from cfMeDIP-seq, H3K4me3 cfChIP-seq, and H3K27ac cfChIP-seq in plasma samples from patients with metastatic PDAC revealed only a moderate correlation between marks **(Figure S3B)**. This contrasts with our observations in the PDX models where the three marks were more highly correlated and may reflect variations in epitope accessibility, epigenomic mark stability, and capture efficiencies of antibodies in plasma vs. tissue. However, since of the 23,944 differential sites across the three epigenomic features, the vast majority (90%; n=21,631) were unique to one epigenomic data type **(Figure S3C)**, the moderate correlation may also suggest that each mark contributes orthogonal information.

### Multianalyte integration improves non-invasive pancreatic cancer subtyping

Compared to tumor tissue RNA-based classical/basal-like labels, plasma epigenomic signal from H3K27ac, H3K4me3, and MeDIP distinguished the two PDAC subtypes with an area under the receiver operating characteristic curve (AUROC) of 0.71, 0.76, and 0.81 **(Figure S4A)**, and area under the precision-recall curve (AUPRC) of 0.79, 0.83, and 0.86, respectively **(Figure S4B)**. Interestingly, cfMeDIP showed numerically higher performance compared to H3K27ac and H3K4me3 individually, which contrasts with our observations in PDX models. This likely reflects the relative stability of DNA methylation as an epigenomic mark compared with histone post-translational modifications leading to its improved recovery in cell-free ctDNA assays. Since each epigenomic feature contributes orthogonal information, we reasoned that integrating these signals may enhance their discriminatory power. Similar to previously described approaches (27), we log-transformed and z-score normalized each individual C:B ratio prior to simple summation to obtain a singular pancreatic integrated epigenomic score, or PIES **(Figure 4A)**. As expected, PIES values were significantly higher in classical compared to basal-like PDAC plasma (p=6.4×10^−8^; **Figure 4B**). To ensure unbiased performance estimates, performance metrics were evaluated using leave-one-out cross-validation (LOO-CV) that yielded an AUROC of 0.84 **(Figure 4C, S4A)** and an AUPRC of 0.89 regardless of ctDNA fraction **(Figure 4D, S4B),** an improvement compared to any individual mark alone. The performance of PIES further improved with an AUROC of 0.9 and AUPRC of 0.92 when focusing on plasma samples with detectable ctDNA levels (>3%, the sensitivity threshold for ichorCNA (50)) **(Figure 4C-D)**. LOO-CV achieved a precision of 88%, a recall of 76%, and a specificity of 84%. The optimal actionable cut-off maximizing the difference between true positive and false positive rates using the Youden index (J), estimated within each training fold and averaged across LOO iterations, was −0.4.

**Figure 4.**
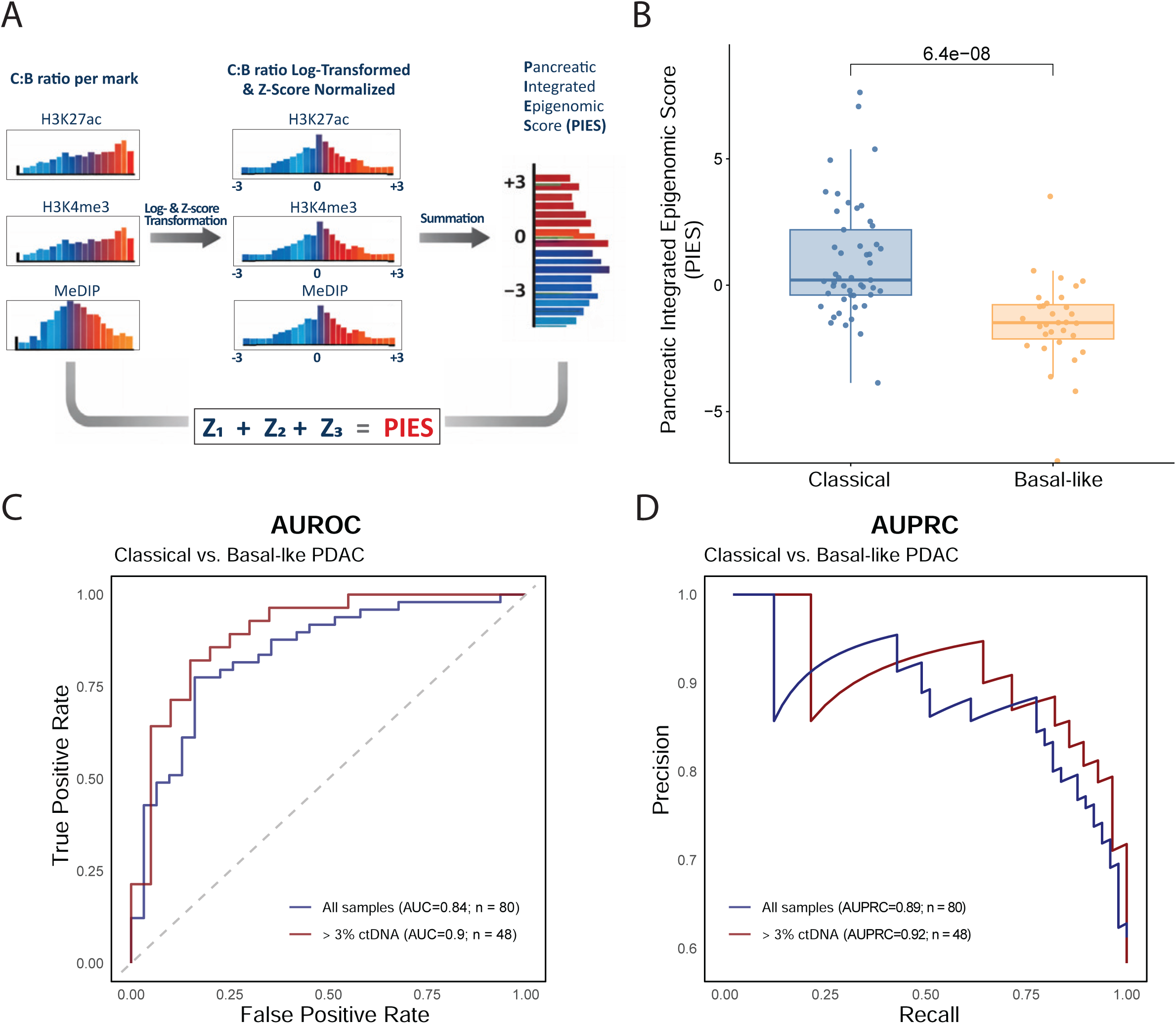
Multianalyte integration improves non-invasive pancreatic cancer subtyping. **(A)** Framework to integrate epigenomic information and evaluate its performance. **(B)** Boxplot comparing pancreatic integrated epigenomic scores for plasma samples of patients with classical and basal-like pancreatic cancer (Wilcoxon rank-sum test). **(C)** Area under the receiver operating characteristic (AUROC) curve for distinguishing classical and basal-like pancreatic cancer via cfDNA epigenomic profiles. **(D)** Area under the precision-recall curve (AUPRC) for distinguishing classical and basal-like pancreatic cancer via cfDNA epigenomic profiles.

### Tumoral subtype heterogeneity may underlie plasma-tissue discordance

The overall accuracy of PIES (compared to tumor tissue RNA labels) was 79% and there was no significant bias in the directionality of inaccurate results (12 out of 49 tissue classical labeled plasma basal-like, and 5 out of 31 tissue basal-like labeled plasma classical subtype; McNemar’s test: p=0.15). Pancreatic ductal adenocarcinoma is a biologically heterogeneous disease, with prior single-cell and spatial profiling studies demonstrating that classical and basal-like transcriptional programs can coexist within the same tumor and even within single malignant cells (5,21). We reasoned that this heterogeneity might partly explain our observed plasma-tissue discordance. To explore this discordance further, we identified 15 cases in our cohort where extra tissue was available from the same site as used for RNA-sequencing and subtype determination. We then applied a validated and orthogonal assay utilizing a six-marker multiplex IF panel which measures quantitative expression levels of classical (GATA6, CLDN18.2, TFF1) and basal-like (KRT5, KRT17, S100A2) proteins in individual tumor cells, categorizing each tumor cell as classical, basal-like or co-expressor as previously described (5) **(Figure 5A)**. Four cases included in this subset were originally discordant between plasma PIES and tissue RNA derived labels (PANFR3025, PANFR3298, and P23 noted to be classical on the plasma PIES and basal-like using tissue RNA; PANFR3130 noted as basal-like by plasma PIES and classical using tissue RNA; **Figure 5B, Table S5**). Interestingly, two of the cases above which were noted as classical by plasma PIES, and basal by tissue RNA (PANFR 3025, and PANFR 3298) were noted as classical on the mIF panel (>50% classical cells on mIF panel as defined previously (5)) but had elements of hybrid co-expressing, and basal-like cells **(Figure 5C)**. On the other hand, PANFR3130 and P23 remained discordant on plasma and tissue testing even after the mIF analysis. PANFR3130 was uniformly classical with 89% classical cells, and P23 was uniformly basal with 92% basal cells on the mIF panel, both concordant with the tissue label determined on bulk RNA-sequencing performed on the same biopsy. Interestingly, and of note, P23 had a PFS of 9 months and an OS of 15 months, substantially higher than expected for a purely basal tumor. Similarly, PANFR3130 exhibited a PFS of 2.3 months and OS of 9.3 months, consistent with a poorer prognosis.

**Figure 5.**
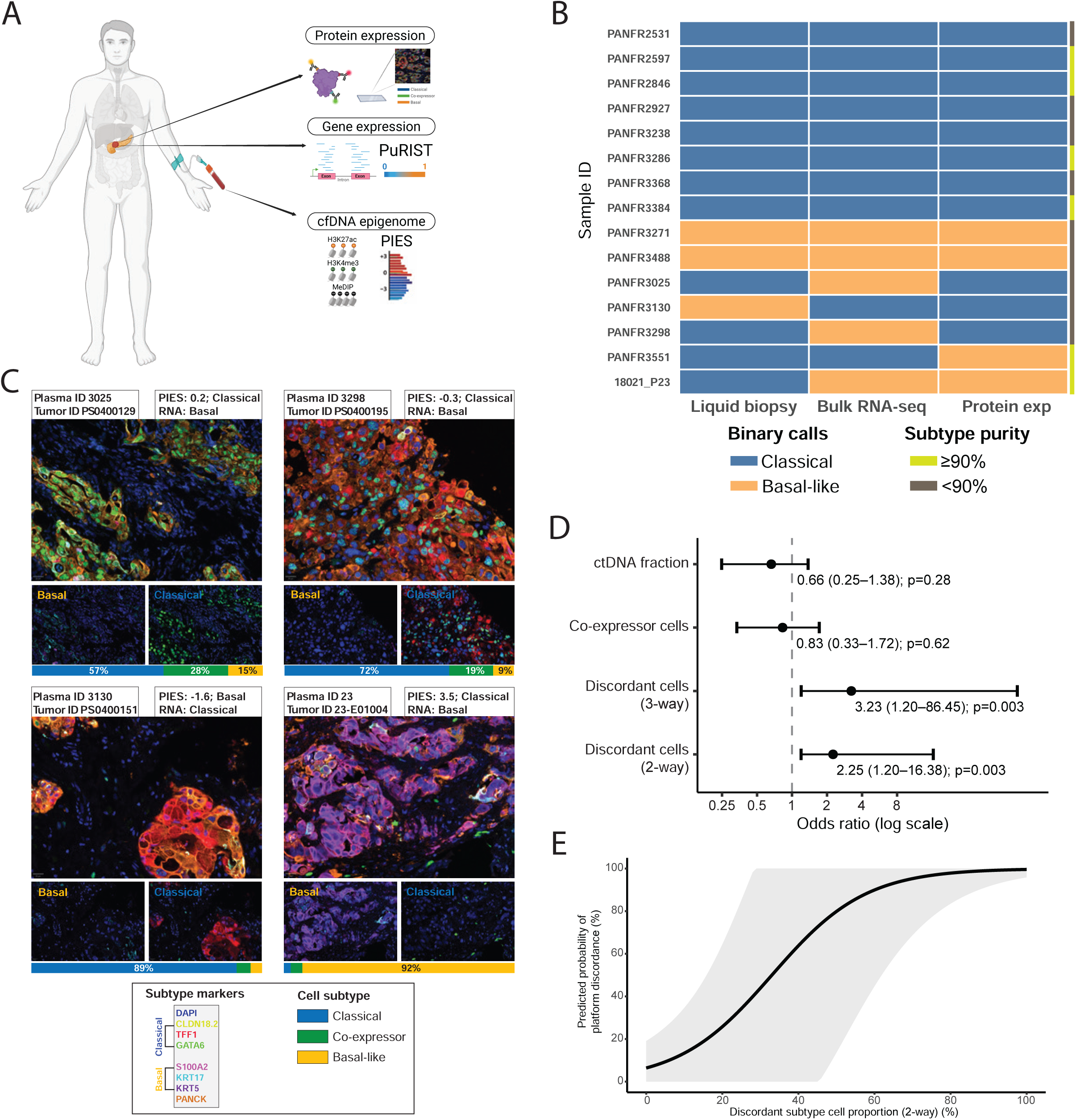
Orthogonal single-cell protein-based expression reveals pancreatic cancer subtype heterogeneity in PDAC and plasma-based subtyping method shows association with prognosis. **(A)** Pancreatic cancer subtyping across single-cell protein-based expression, bulk RNA-seq, and cfDNA epigenomic profiling. **(B)** Comparison of subtype calls across three subtyping platforms: liquid biopsy epigenomic profiles, bulk RNA-seq, and single-cell protein-based expression. **(C)** Representative multiplex immunofluorescence (mIF) images from four pancreatic cancer tumors. Scale bar = 20 μm. **(D)** Forest plot of univariate logistic regression analyses assessing the association between platform discordance and biological variables, including circulating tumor DNA (ctDNA) fraction, co-expressor cell proportion, and discordant subtype cell proportion derived from two-group and three-group classification in mIF. Odds ratios are reported per 10 percentage-point increase. Bars represent the 95% CI. **(E)** Marginal effects analysis showing increased PIES-discordant subtype proportion in mIF is associated with increased discordance across tissue and plasma-based subtyping platforms.

To understand discordance more systematically, we conducted univariate logistic regression to identify predictors of plasma PIES and tissue subtype discordance. Biological variables such as ctDNA tumor fraction, the proportion of co-expressor cells, and the proportion of PIES-discordant subtype cells (i.e. frequency of cell type discordant to plasma PIES determination on tumor mIF panel analysis) were included as independent variables. Interestingly, increasing representation of the PIES-discordant subtype on mIF panel analysis by two-group (i.e. only counting classical and basal cells as denominator to assess discordant cell type frequency) and three-group classifications (counts classical, basal and co-expressor cells as denominator to assess discordant cell type frequency) were associated with an increased likelihood of discordance across platforms (two-group odds ratio per 10% increase [OR]=2.25, 95% CI: 1.2-16, p=0.003; three-group OR=3.23, 95% CI: 1.2-86, p=0.003; respectively; **Figure 5D**). ctDNA tumor fraction and the proportion of co-expressor cells by mIF were not associated with an increased likelihood of discordance between plasma and tissue-based subtyping platforms (OR=0.66, 95% CI: 0.25-1.38, p=0.28; OR=0.83, 95% CI: 0.33-1.72, p=0.62; respectively). Marginal effects analysis demonstrated a monotonic increase in the predicted probability of discordance across the observed range of discordant cell type in tumors. Tumors with low proportions of PIES-discordant cells exhibited a low predicted probability of discordance, whereas tumors in which the discordant subtype predominated showed a markedly higher predicted probability **(Figure 5E)**. These findings indicate that increasing dominance of the PIES-discordant subtype is associated with progressively greater discordance across classification platforms, whereas more homogeneous tumors are more likely to be concordant between tissue and blood-based approaches. Interestingly, mixed tumors, previously defined as those with <90% of classical or basal-like cells on mIF (5) did not have intermediate-range PIES scores compared to more pure tumors **(Figure S3B)**, and hence the score threshold alone may not allow identification of such cases.

### Plasma subtypes improve clinical outcome prediction compared to tumor tissue subtyping

Using the findings above, we hypothesized that plasma-based subtype determination may better predict clinical outcomes. For this, we analyzed survival data from patients who had plasma collected prior to the initiation of the first-line treatment for pancreatic cancer (n=50). The median (IQR) age was 65 (58—70), 31 were male (62%), 35 (70%) had liver metastases, and 32 (64%) had ECOG greater than or equal to 1 **(Figure 6A; Table S4**). The first-line regimens administered were FOLFIRINOX (FFX; n=17) or Gemcitabine + nab-paclitaxel (GnP; n=33). We categorized patients into two groups: low and high PIES using a cut-off of −0.4 derived using the Youden index (J) across all LOO iterations. In a multivariable Cox regression model accounting for ctDNA fraction (continuous), the presence vs. absence of liver metastasis, ECOG (0 vs. 1-2), CA19-9 levels (continuous), and the chemotherapy regimen administered (FFX vs. GnP), low PIES (vs. high PIES) but not tissue transcriptomic subtype (basal-like vs. classical) emerged as an independent determinant of worse progression-free survival (PFS) in patients with metastatic PDAC (Low PIES: adjusted HR=2.99; 95% CI: 1.3—6.9; p=0.01; **Figure 6B**; basal-like tissue RNA subtype: adjusted HR=2; 95% CI: 0.94—4.24; p=0.07; **Figure S6A**). Patients with low PIES had a significantly worse PFS compared to patients with high PIES, with a median (95% CI) PFS of 4.7 (2.3—NA) vs. 6.9 (4.8—11.2) months (log-rank p=0.046; **Figure 6C**), and, similar to the results of the multivariable Cox analysis, PFS among classical and basal-like PDAC determined using tissue-based transcriptomic analysis did not reach statistical significance (5.4 vs. 6.4 months; log-rank p=0.25; **Figure 6D**).

**Figure 6.**
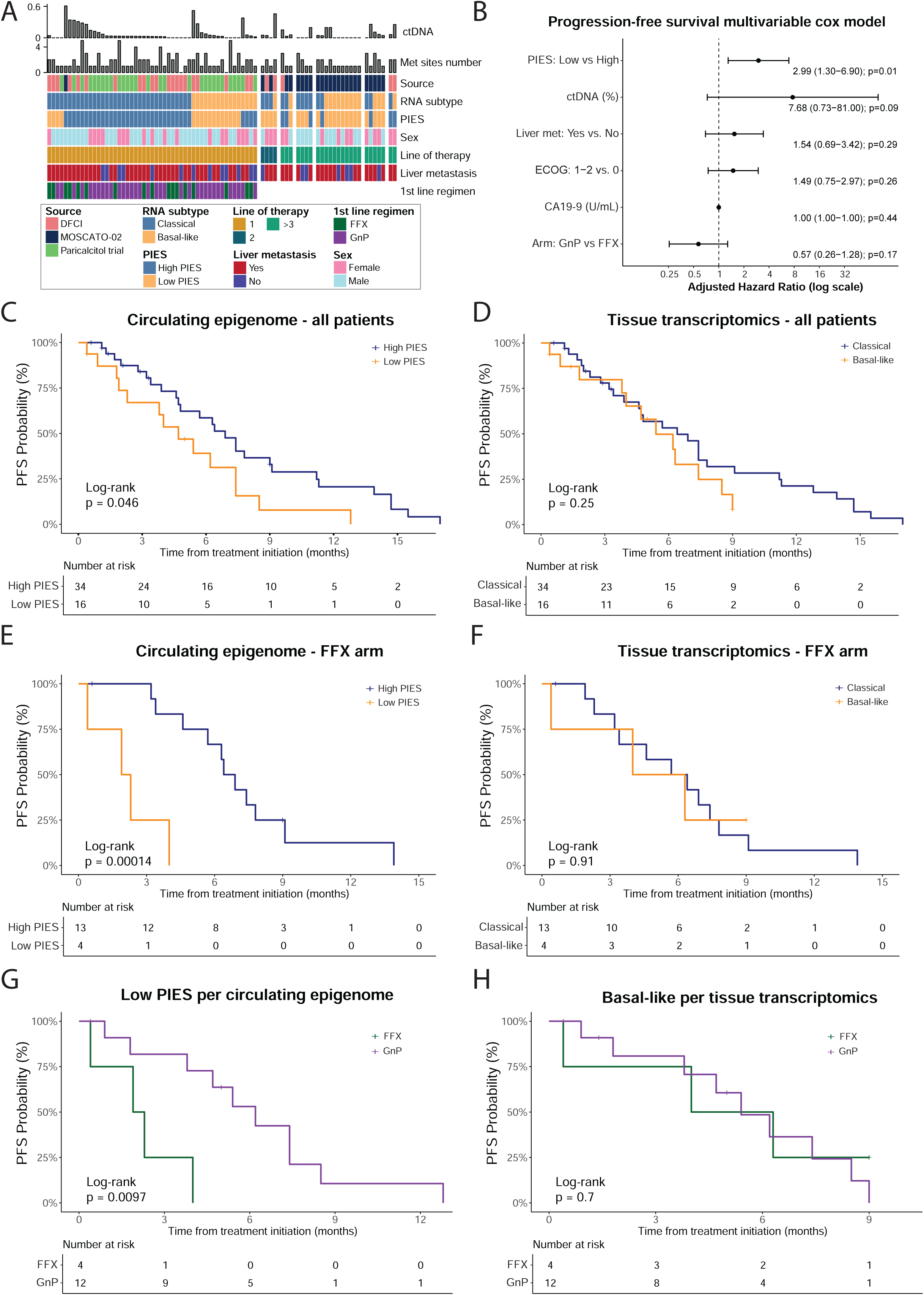
Clinical outcomes stratified by circulating epigenomic and tissue transcriptomic PDAC subtypes across treatment regimens. **(A)** Demographics and clinical characteristics of patients in the PDAC plasma cohort **(B)** Forest plot for multivariable Cox proportional hazards regression model for progression-free survival of patients with metastatic PDAC. Bars represent the 95% CI. **(C)** Kaplan-Meier curves of progression-free survival of patients with metastatic PDAC stratified by high and low PIES. **(D)** Kaplan-Meier curves of progression-free survival of patients with metastatic PDAC stratified by tissue transcriptomics subtypes. **(E)** Kaplan-Meier curves of progression-free survival of patients with metastatic PDAC receiving FOLFIRINOX (FFX) stratified by high and low PIES. **(F)** Kaplan-Meier curves of progression-free survival of patients with metastatic PDAC receiving FFX stratified by tissue transcriptomics subtypes. **(G)** Kaplan-Meier curves of progression-free survival of patients with metastatic PDAC with low PIES stratified by treatment regimen. **(H)** Kaplan-Meier curves of progression-free survival of patients with metastatic PDAC with basal-like subtype per tissue transcriptomics stratified by treatment regimen.

Given prior evidence that classical and basal-like PDAC exhibit differential responses to FFX and GnP (15,13), with basal-like tumors faring poorly on FFX compared to GnP, we performed a sensitivity analysis stratified by treatment regimen to assess whether the prognostic impact of PIES varied across therapeutic subgroups. Interestingly, among patients treated with FFX, patients with low PIES had a more pronounced poor prognosis compared to patients with high PIES, with a median (95% CI) PFS of 2.1 (0.4—NA) vs. 6.7 (5.7—NA) months (p<0.001; **Figure 6E**). In contrast, we did not observe differences in outcomes among patients with PDAC receiving FFX according to tissue-based transcriptomic analysis with a median (95% CI) PFS of 5.2 (0.4—NA) vs. 6.1 (3.4—NA) months (p=0.91; **Figure 6F**). These differences were not observed among patients treated with GnP **(Figure S6B-C)**. Moreover, when stratified by treatment arms (FFX vs. GnP), among patients with low PIES, patients treated with GnP had better outcomes compared to patients treated with FFX, with a median (95% CI) PFS of 6.2 (4.7—NA) vs. 2.1 (0.4—NA) months (p<0.01; **Figure 6G**). Conversely, we did not observe differences in outcomes among regimens in patients with a basal-like subtype per tissue transcriptomics (p=0.7; **Figure 6H**). Finally, we included an interaction term between chemotherapy regimen (FFX vs. GnP) and PIES group (high vs. low) in the Cox regression model for PFS, adjusting for ctDNA fraction and the presence of liver metastases. A significant interaction between chemotherapy regimen and PIES group was observed (interaction p=0.04), with patients with low PIES appearing to derive greater benefit from GnP compared with FFX. Overall, these results suggest that PIES may better capture underlying tumor biology and be more representative of cancer behavior than tissue transcriptomics, which can be limited by PDAC intratumoral and intermetastatic heterogeneity.

## Discussion

In this study, we introduce the Pancreatic Integrated Epigenomic Score (PIES), a novel non-invasive approach for inferring pancreatic cancer transcriptional subtypes based on circulating tumor epigenomic profiles. Leveraging a large PDAC PDX cohort, we identified highly recurrent gene regulatory programs that distinguish classical and basal-like subtypes and validated these findings in an independent PDAC PDX cohort. Subsequently, we capture subtype-specific epigenomic signals in the plasma of patients with advanced metastatic PDAC and develop PIES by integrating three distinct epigenomic analytes. We show that PIES accurately discriminates between classical and basal-like PDAC in patients with metastatic disease, with low PIES indicating a basal-like subtype and high PIES indicating a classical subtype. Finally, we show that PIES improves prognostication and treatment response prediction over tissue-based RNA subtyping by providing a readout of the overall tumoral transcriptional subtype that better accounts for prevalent intratumoral and intermetastatic lesion subtype heterogeneity.

Efforts to subtype pancreatic cancer in the clinic have been limited in practice by the need for invasive tissue biopsy, low tumor cellularity of pancreatic tumors, leaving insufficient tumor for testing, and biologically by marked spatial tumor heterogeneity within a single lesion and across different metastatic sites (5,13,15,54). By capturing tumor-derived material shed from multiple sites (55,56), our approach provides a promising non-invasive alternative that reflects the overall tumor burden subtype, rather than focusing on a single lesion or being constrained by limited tissue availability. Moreover, its non-invasive nature enables longitudinal assessment, offering the potential to monitor subtype shifts in response to treatment, which have been reported in approximately 20% of PDAC cases (8). On the other hand, liquid biopsy approaches are often limited by the amount of ctDNA shed into the bloodstream, and pancreatic cancer is known to be a low-ctDNA-shedding tumor (51,52), a finding that was also seen in our study. Nevertheless, PIES performed well across our cohort, albeit slightly improved at tumor fractions above 3%. This likely stems from the integrative nature of our approach, which relies on the aggregate of tumor signals across multiple genomic loci and incorporates three complementary epigenomic marks that contain orthogonal information.

A major strength of this study is the breadth and depth of epigenomic profiling across a large PDAC PDX cohort and that we employed two validated and widely used RNA-based subtyping algorithms, PurIST and Moffitt, to determine their transcriptional subtypes (7,9). We identified highly recurrent gene regulatory programs that distinguish classical from basal-like PDAC PDXs. Leveraging these programs, we accurately inferred the transcriptional subtype of PDAC PDXs in an independent external cohort, reinforcing the hypothesis that these subtypes can be inferred from the epigenome (39). We then queried the signal at subtype-specific genomic loci using the same epigenomic assays in the plasma of patients with advanced pancreatic cancer, capturing PDAC transcriptional subtype through minimally invasive sampling. Our plasma cohort included real-world biobanked samples at an academic institution and prospectively collected plasma samples from two clinical trials conducted in Europe and the United States for patients treated in varied clinical settings which support the generalizability of our findings.

In our study, PIES was associated with clinical outcomes in patients with PDAC. Patients with low PIES had significantly worse outcomes than those with high PIES when treated with FFX, whereas no difference in outcomes was observed among patients treated with GnP. Interestingly, among patients with low PIES, those treated with GnP had significantly better outcomes than patients treated with FFX. This finding supports findings of the PASS-01 (NCT04469556) and COMPASS trials (NCT02750657) that patients with basal-like PDAC may derive better outcomes with GnP (13,15). In contrast, there was no significant difference in outcomes among patients with PDAC when stratified by transcriptional subtypes based on tissue biopsy. While several studies reported a statistically significant worse prognosis for patients with basal-like compared to classical PDAC (8,15,16), others have observed only numerical differences that did not reach statistical significance (13). Similarly, while some studies have suggested that basal-like PDAC may derive greater benefit from GnP compared with FFX(13,15), others have been unable to reproduce these findings (8). Such discrepancies may, in part, reflect the impact of intratumoral and intermetastatic lesion heterogeneity, which may have limited our understanding of the clinical impact of transcriptional subtypes. As novel therapeutic approaches continue to emerge (18,57), the need for robust assays capable of overcoming spatial heterogeneity becomes increasingly critical to advance the standard of care in PDAC.

We acknowledge important limitations of the study. First, although PDX models offer the advantage of relatively high tumor purity and larger tumor volumes, we acknowledge they may not be fully representative of the tumor biology and microenvironment. Second, the ground truth labels for the plasma samples were derived using PurIST in two out of three clinical cohorts and spatial transcriptomic profiling using the GeomX Digital Spatial Profiler in the third cohort (NCT03520790). Third, although our findings were validated across independent PDX cohorts and three plasma cohorts, the plasma sample size remains modest, and larger multi-institutional studies will be necessary to establish standardized thresholds for subtype determination prior to clinical implementation. Lastly, we acknowledge that cell-free DNA fragmentation patterns, nucleosome positioning, and capture efficiencies differ from those observed in tissue chromatin, which may have influenced the performance of our subtype inference model that was based on tissue-informed signatures. Our current plasma assays do not provide the signal-to-noise ratio to develop plasma-informed signatures without any tissue-based genomic loci restriction, though in the future such approaches may allow a more refined and granular view on in vivo subtypes in PDAC.

In conclusion, our study highlights the emerging potential of liquid biopsy and cell-free DNA epigenomic profiling as a novel circulating biomarker for PDAC subtyping to inform prognostication and therapeutic decision-making. This approach requires only 1 mL of plasma and therefore is feasible for broad and rapid clinical implementation. By enabling accurate molecular subtyping, it may facilitate subtype-informed clinical trial enrollment, advance the development of tailored therapeutic strategies, and support more precise treatment decision-making in pancreatic cancer.

## Methods

### Patient-derived xenografts

Patient-derived xenografts (PDX) were derived from patients with pancreatic ductal adenocarcinoma (PDAC) from two institutions: Dana-Farber Cancer Institute (DFCI) and MD Anderson Cancer Center (MDACC). Experiments were conducted under approved protocol at Dana-Farber Cancer Institute and conducted in accordance with Institutional Animal Care and Use Committee (IACUC, protocol 14-030). Tumors were established by subcutaneous implantation of patient tumor samples and expanded through serial transplantation in mice. Briefly, tumors were collected from patients, sectioned into small fragments (2 mm^3^), and implanted subcutaneously into 6–8-week-old female nude mice (Charles River Laboratories). Tumor growth was monitored, and when tumors reached around 2cm, they were excised, fragmented, and re-implanted. This process was repeated to expand the model in vivo across multiple generations. PDX models provided by MDACC were developed and established as previously described (58) by implanting tissue fragments at 4 mm^3^ fragment volume subcutaneously into the right flank of 6-8 week old NSG mice. Tumor volumes were captured by serial caliper measurements weekly. Tumor volume (TV) was calculated as TV = (D × d2/2), where “D” is the larger and “d” is the smaller superficial visible diameter of the tumor mass. All measurements were documented as mm^3^. At the endpoint of the study, tumors were collected and fixed in 10% neutral buffered formalin overnight and then processed and embedded in paraffin for histology analysis or snap frozen in liquid nitrogen for sequencing analysis.

### Plasma samples acquisition

Plasma samples from patients with metastatic pancreatic cancer were obtained from (1) a randomized phase I/II trial (NCT03520790), (2) a prospective nonrandomized clinical trial MOSCATO-02 (NCT01566019), (3) a collection of plasma samples from patients treated at DFCI under IRB-approved protocol (DFCI 03-189). All patients provided written informed consent. Studies were conducted in accordance with recognized ethical guidelines. IRB approvals for the NCT03520790 and MOSCATO-02 (NCT01566019) trials are described in the original publications.

### Subtype calling of tumors in plasma clinical cohorts

Tumor samples obtained from the MOSCATO-02 (NCT01566019) trial and from patients with pancreatic cancer treated at the Dana-Farber underwent profiling by bulk RNA-seq. Classical and basal-like transcriptional subtypes were determined using the PurIST algorithm (9).Tumor samples obtained from a Phase I/II clinical trial evaluating the safety of the vitamin D receptor agonist paricalcitol in combination with gemcitabine and nab-paclitaxel in patients with metastatic pancreatic ductal adenocarcinoma (NCT03520790) underwent profiling by the NanoString GeoMx Digital Spatial Profiler (DSP) Whole Transcriptome Assay as previously described (Dias Costa & Perez et al). After quality control and batch correction, the resulting gene expression data from tumor centers were scored for the expression of the scClassical and scBasal gene signatures via the single-sample GSEA algorithm as implemented by the GSVA package (Dias Costa & Perez et al 2026)(21). Scores were then averaged at the tumor level, and each tumor was classified as primarily of the classical or basal subtype depending on which signature score was highest.

### PDX tissue chromatin immunoprecipitation followed by sequencing (ChIP-seq)

Frozen tissue was pulverized using the Covaris CryoPrep system and fixed with 2 mmol/L disuccinimidyl glutarate for 10 minutes followed by 1% formaldehyde for 10 minutes and quenched with glycine. Chromatin was sheared using the Covaris E220 ultrasonicator and then incubated overnight with the following antibodies coupled with 40 μL protein A and protein G beads (Invitrogen) at 4°C overnight: H3K27ac (Abcam #ab4729), H3K4me3 (Thermo Fisher Scientific #PA5-27029). Five percent of the sample was not exposed to antibody and was used as a control “input”. Beads were washed three times each with Low-Salt Wash Buffer (0.1% SDS, 1% Triton X-100, 2 mmol/L EDTA, 20 mmol/L Tris-HCl pH 7.5, 150 mmol/L NaCl), High-Salt Wash Buffer (0.1% SDS, 1% Triton X-100, 2 mmol/L EDTA, 20 mmol/L Tris-HCl pH 7.5, 500 mmol/L NaCl), and LiCl Wash Buffer (10 mmol/L Tris pH 7.5, 250 mmol/L LiCl, 1% NP-40, 1% Na-Doc, 1 mmol/L EDTA) and rinsed with TE buffer (pH 8.0) once. Samples were then de-cross-linked, treated with RNase and proteinase K, and DNA was extracted using MinElute PCR Purification Kit (Qiagen). DNA sequencing libraries were prepared from the purified immunoprecipitated and input DNA using the ThruPLEX DNA-seq Kit (TakaraBio). Libraries were sequenced on the Illumina NovaSeq X Plus platform to generate 150 bp paired-end reads (Novogene Corporation).

### PDX tissue assay for transposase-accessible chromatin sequencing (ATAC-seq)

Frozen tissue was resuspended and dounce homogenized in 1,000 μL of homogenization buffer. Nuclei were filtered using a 70-μm Flowmi strainer, isolated using iodixanol density-gradient centrifugation method, and washed with RSB buffer (10 mmol/L Tris-HCl pH 7.4, 10 mmol/L NaCl, and 3 mmol/L MgCl2 in water). Fifty thousand nuclei were resuspended in 50 μL of transposition mix [2.5 μL transposase (100 nmol/L), 16.5 μL PBS, 0.5 μL 1% digitonin, 0.5 μL 10% Tween-20, and 5 μL water]. Transposition reactions were incubated at 37°C for 30 minutes in a thermomixer shaking at 1,000 rpm. Reactions were cleaned with Qiagen columns. Libraries were amplified using the Omni-ATAC protocol and sequenced on the Illumina NovaSeq X Plus platform (Novogene Corporation) using 150-base paired-end reads (59,60).

### Methylated DNA immunoprecipitation followed by sequencing (MeDIP-seq)

Methylated DNA immunoprecipitation sequencing (MeDIP-seq) was performed on tissue and plasma following published methods(25,49,61,62). Library preparation was performed on 10 ng of DNA using the KAPA HyperPrep Kit (KAPA Biosystems). We then performed end-repair, A-tailing, and ligation of NEBNext adaptors (NEBNext Multiplex Oligos for Illumina kit, New England BioLabs). Libraries were digested using the USER enzyme (New England BioLabs). λ DNA, consisting of unmethylated and *in vitro* methylated DNA, was added to prepared libraries to achieve a total amount of 100 ng DNA. Methylated and unmethylated *Arabidopsis thaliana* DNA (Diagenode) was added for quality control. MeDIP was performed using the MagMeDIP Kit (Diagenode) following the manufacturer’s protocol. Samples were purified using the iPure Kit v2 (Diagenode). Success of the immunoprecipitation was confirmed using qPCR to detect recovery of the spiked-in *Arabidopsis thaliana* methylated and unmethylated DNA. KAPA HiFi Hotstart ReadyMix (KAPA Biosystems) and NEBNext Multiplex Oligos for Illumina (New England Biolabs) were added to a final concentration of 0.3 μmol/L and libraries were amplified. Samples were pooled and sequenced on the Illumina NovaSeq X Plus platform (Novogene Corporation) to generate 150 bp paired end reads.

### Patient-derived xenografts RNA-seq and transcriptional subtype calling

RNA was extracted from frozen tumor samples using the Qiagen RNeasy Mini Kit (Cat No./ID: 74104). RNA sequencing (RNA-seq) libraries were constructed from 1 μg RNA using the Illumina TruSeq Stranded mRNA LT Sample Prep Kit. Barcoded libraries were pooled and sequenced on the Illumina Novaseq X plus platform. FASTQ files were processed using the VIPER workflow(63). Read alignment to human genome build hg19 was performed with STAR (RRID:SCR_004463)(64). Cufflinks (RRID:SCR_014597) was used to assemble transcript-level expression data from filtered alignments (65). Classical and basal transcriptional subtypes were identified using bulk RNA-seq and the purity-independent subtyping (PurIST) algorithm (9). Briefly, PurIST is a single-sample classifier that compares the expression of eight gene pairs to develop a probability of classical and basal tumors. PurIST classifications were also compared with Moffitt subtype assignments and showed complete concordance across all 28 PDX models used in the subsequent analyses (7).

### Cell-free chromatin immunoprecipitation followed by sequencing (cfChIP-seq)

Twenty-five micrograms of antibody were conjugated to 5mg of Dynabeads M-270 Epoxy (Invitrogen, cat # 14311D) according to the manufacturer’s instructions. After coupling, beads were stored at 4 °C and pre-cleared in 0.1% BSA in PBS for 5 min at 4 °C prior to use. The following antibodies were used: H3K27ac (Abcam #ab4729) and H3K4me3 (Thermo Fisher Scientific #711958). 810 µl of thawed plasma was supplemented with protease inhibitor cocktail (Roche, 11873580001), centrifuged at 3,000g for 15 min at 4 °C, and combined with 90 µl of salt–detergent. The resulting 900uL of plasma was incubated with 1ug of antibody-coupled magnetic beads at 4°C with rotation. Beads were washed three times each with Low-Salt Wash Buffer (0.1% SDS, 1% Triton X-100, 2 mmol/L EDTA, 150 mmol/L NaCl, 20 mmol/L Tris-HCl pH 7.5), High-Salt Wash Buffer (0.1% SDS, 1% Triton X-100, 2 mmol/L EDTA, 500 mmol/L NaCl, 20 mmol/L Tris-HCl pH 7.5), and LiCl Wash Buffer (250 mmol/L LiCl, 1% NP-40, 1% Na Deoxycholate, 1 mmol/L EDTA, 10 mmol/L Tris-HCl pH 7.5) and rinsed with TE buffer once (Thermo Fisher Scientific, cat #BP2473500). Subsequently, beads were resuspended and incubated in 100 μL of DNA extraction buffer containing 0.1 mol/L NaHCO3, 1% SDS, and 0.6 mg/mL Proteinase K (Qiagen, cat #19131) and 0.4 mg/mL RNaseA (Thermo Fisher Scientific, cat #12091021) for 10 minutes at 37°C, for 1 hour at 50°C and for 2 hours at 65°C. DNA was purified using the ChIP DNA Clean & Concentrator Kit (Zymo Research, cat #D5205). Cell-free ChIP-seq (cfChIP-seq) libraries were prepared with ThruPLEX DNA-Seq Kit (Takara Bio, cat #R400675) following the manufacturer’s instructions. After library amplification, the DNA was purified by AMPure XP (Beckman coulter, cat# A63880). The library was submitted for the 150 base-pair paired-end sequencing on the Illumina NovaSeq X Plus platform (Novogene Corporation).

### Cell-free DNA extraction and circulating tumor DNA estimation in plasma samples

Cell-free (cf)DNA extraction and low-pass whole-genome sequencing (LPWGS) were performed on plasma samples using previously published methods (23). LPWGS reads were aligned using the SNAPIE (Streamlined Nextflow Analysis Pipeline for Immunoprecipitation-based Epigenomics) with default parameters (66). The ichorCNA R package (RRID:SCR_024768) was used to infer copy-number profiles and cfDNA tumor content from read abundance across bins spanning the genome using default parameters (50).

### ChIP-seq, ATAC-seq and MeDIP-seq peak calling

Chromatin immunoprecipitation sequencing (ChIP-seq) and Assay for Transposase-Accessible Chromatin using sequencing (ATAC-seq) reads were aligned to the human genome build hg19 using the Burrows-Wheeler Aligner version 0.7.17 (RRID:SCR_010910) (67). Methylated CpG dinucleotide immunoprecipitation and sequencing (MeDIP-seq) reads were aligned with the methyl-seq pipeline v3.0.0 from the nf-core repository (68) (https://nf-co.re/). Non-uniquely mapped and redundant reads were discarded. MACS v2.1.1.20140616 (RRID:SCR_013291) was used for ChIP-seq, ATAC-seq, and MeDIP-seq peak calling with a q-value threshold of 0.01. IGV v2.8.2 was used to visualize normalized ChIP-seq read counts at specific genomic loci (69,70). ChIP-seq heatmaps were generated with deepTools v3.3.1 (RRID:SCR_016366) and show normalized read counts at the peak center ±2 kb unless otherwise noted (71). Overlap of ChIP-seq peaks was assessed using BEDTools v2.26.0. Peaks were considered overlapping if they shared one or more base pairs. For plasma data including cfChIP-seq profiling of histone modifications (H3K4me3 and H3K27ac), cfMeDIP-seq, low-pass whole-genome sequencing we used the SNAPIE (Streamlined Nextflow Analysis Pipeline for Immunoprecipitation-based Epigenomics) with default parameters (66).

### Defining sets of genomic loci to distinguish classical vs. basal-like PDAC

H3K27ac and H3K4me3 ChIP-seq, and ATAC-seq from classical and basal-like PDAC PDX were compared to identify genomic loci (peaks) with significant enrichment in the two above groups referred to as “classical-up” and “basal-up” peaks. A union set of peaks was created using BEDTools v2.26.0, and narrowPeak calls from MACS were used for H3K27ac, H3K4me3 and ATAC. The number of unique aligned reads overlapping each genomic locus (peak) in each sample is calculated from BAM files using BEDtools v2.26.0, to create a count matrix in which each row represents a peak, and each column represents a sample. Read counts for each peak were normalized to the total number of mapped reads for each sample. Using DEseq2 v1.44.0 (41), subtype-enriched peaks were identified at a defined false discovery rate (FDR)-q value and log_2_ fold-change (LFC) thresholds (for H3K27ac and ATAC: FDR-q < 0.05, |LFC| > 0; for H3K4me3: FDR-q < 0.05, |LFC| > 0). Unsupervised hierarchical clustering was performed based on Spearman correlation between samples. Principal component analysis was performed using the prcomp R function v4.4.0. The GREAT analysis v4.0.4 was used to assess enrichment of Gene Ontology (GO) annotations among genes near differential ChIP-seq peaks, assigning each peak to the nearest gene within 500 kb (72).

### Refining enhancer-centric (ATAC and H3K27ac) epigenomic signature

Sites that were both differentially marked by H3K27ac and differentially accessible were classified as “intersection” sites. Their performance was compared to regions that were exclusively differentially accessible (“ATAC-only”), exclusively differentially marked by H3K27ac (“H3K27ac-only”), as well as the complete sets of differential ATAC peaks and differential H3K27ac peaks. For each site set, we computed an aggregate signal per sample by summing the values across all sites and normalizing by the number of sites in the set. To quantify subtype separation, we performed exhaustive pairwise comparisons between all classical and basal-like samples. For each classical-basal sample pair, we calculated the absolute difference in normalized aggregate signal, generating a distribution of pairwise subtype separation values for each site set. The mean separation across these pairwise comparisons was then used to evaluate the discriminatory capacity of each region category using a Wilcoxon signed-rank test.

### Refining of promoter-centric (H3K4me3) epigenomic signature

Promoter regions were defined across a range of log fold-change (LFC) thresholds, and for each threshold we calculated the per-sample normalized aggregate signal across selected promoters. Similar to the framework described above, subtype separation was then quantified as the absolute difference in normalized signal across all pairwise combinations of classical and basal-like samples. For each classical-basal sample pair, we calculated the absolute difference in normalized aggregate signal, generating a distribution of pairwise subtype separation values for each site set. The mean separation across these pairwise comparisons was then used to evaluate the discriminatory capacity of each region category using a Wilcoxon signed-rank test.

### Generation of the pancreatic integrated epigenomic score (PIES) for plasma samples

For cfChIP-seq (both H3K4me3 and H3K27ac) and cfMeDIP-seq, and for each plasma sample, we counted reads overlapping with the previously defined “classical-up” and “basal-up” peaks. For each epigenomic mark, and for each plasma sample, a ratio of reads mapping at the “classical-up” sites over reads mapping at the “basal-up” sites was calculated. To generate the pancreatic integrated epigenomic score, we log-transformed and Z-score normalized each of the 3 ratio and summed them, for each plasma sample. Since “classical-up” reads are the numerator of the ratio and “basal-up” reads are the denominator, a higher integrated epigenomic score indicates a higher likelihood of categorizing the plasma sample as classical PDAC. Conversely, a lower score suggests a higher likelihood of categorizing the sample as basal-like PDAC. Finally, we characterized the ability of the PIES score to recover classical vs. basal labels by measuring the area under the receiver operating characteristic (AUROC) curve and area under the precision-recall curve (AUPRC).

### Leave-one-out cross-validation

Leave-one-out cross-validation (LOO-CV) was performed on all available plasma samples, consisting of 49 classical and 31 basal-like samples (total n=80). For each iteration, the held-out sample was excluded from all preprocessing steps. For each iteration, one plasma sample was left out, and Youden’s index (True positive rate – False positive rate) was calculated on the remaining 79 samples to identify the optimal threshold that maximized the index. Normalization parameters used to compute the PIES were estimated exclusively from the training samples in each fold and subsequently applied to the held-out sample to generate an out-of-fold prediction. This threshold was then applied to the PIES score of the left-out sample to assign a prediction: samples with scores above the threshold were classified as classical, and those below as basal-like. After all 80 samples were individually evaluated, the PIES labels were compared to the RNA subtype labels, and performance metrics, including recall (sensitivity), precision, and specificity, were calculated.

### Data Analysis and Statistics

Unless differently stated, all pairwise tests are unpaired and two-tailed Wilcoxon rank sum tests. All reported boxplots are based on first quartile, median, and third quartile, with whiskers extending up to 1.5 IQR. Unless otherwise stated, all correlation coefficients are Spearman’s R rounded to the second digit. Survival data was extracted from the MOSCTAO02 trial (NCT01566019), a trial evaluating the use of high throughput molecular analysis to treat patients with metastatic cancer with targeted therapeutics, a randomized phase I/II study evaluating the use paricalcitol plus gemcitabine and nab-paclitaxel in patients with metastatic pancreatic cancer (NCT03520790), and collected retrospectively for patients treated at Dana-Farber Cancer Institute. Kaplan–Meier curves and log-rank P values were computed and plotted using the survival v3.5.8 and survminer v0.4.9 R packages. Reported analyses and plots were created with R 4.4.0 with tidyverse packages. Given the retrospective nature of this study, no randomization was applied. No power analysis was conducted, and size was dictated by sample availability.

### Multiplex immunofluorescence (mIF)

Multiplex immunofluorescence (mIF) was performed using a previously developed and validated tumor epithelial subtype panel, as described (5). Briefly, 4 µm FFPE tissue sections were stained with a six-marker epithelial subtype panel comprising basal markers (KRT17, KRT5, S100A2) and classical markers (GATA6, TFF1, CLDN18.2), together with DAPI for nuclear identification and pan-cytokeratin for epithelial cell identification. Multiplex staining was performed on a Leica BOND RX Research Stainer (Leica Biosystems, Buffalo, IL), and imaging was conducted using the Vectra Polaris system (PhenoImager HT) (Akoya Biosciences, Waltham, MA). Multispectral images were unmixed and analyzed using machine-learning-based tissue and cell segmentation in inForm software (inForm 3.0, Akoya Biosciences), followed by single-cell-level data export and downstream analysis in R (v4.3.2, R Foundation for Statistical Computing; Vienna, Austria).

## Data availability

Processed data are available in the Gene Expression Omnibus (GEO) under accession number GSE326059 (reviewer token: qtahyssmxxavpyf). Raw human sequencing data generated and analyzed in this study are being deposited in dbGaP under controlled access to protect participant privacy (accession pending). These datasets include (1) tissue ChIP-seq profiling of histone modifications (H3K4me3, and H3K27ac), ATAC-seq, MeDIP-seq, and RNA-seq for all PDX models, (2) plasma cfChIP-seq profiling of histone modifications (H3K4me3, and H3K27ac), cfMeDIP-seq, and low-pass whole-genome sequencing.

## Code availability

Scripts to reproduce analyses from this study are available at (https://github.com/Baca-Lab/scripts-cfChIP-PDAC).

## Authors’ Disclosures

**K.S., M.E., A.J.A, S.C.B., B.M.W., M.L.F. and H.S.** have a patent application related to methods for subtyping pancreatic cancer through cfDNA epigenomic analysis (US 63/777,458). **G.S.G.** is a co-founder and shareholder of CytoTRACE Biosciences. **J.E.B.** reports receiving speaker honoraria from Guardant Health; serving in a consulting/advisory role for Guardant Health, Precede Biosciences, Genome Medical, Oncotect, Tracer Biotechnologies, and Musculo; holding equity in Precede Biosciences, Genome Medical, Oncotect, Tracer Biotechnologies, Musculo, and Cityblock Health; and receiving research funding from Guardant Health and Precede Biosciences. **S.C.B., M.L.F., and T.K.C.** are co-founders and shareholders of Precede Biosciences. **B.H.** reports consulting role for Lilly, and research funding to institution from Bristol Myers Squibb. **J.M.C.** receives research funding to his institution from Amgen, Artios, Partners Therapeutics, Roche, Servier, and Bristol Myers Squibb. He receives research support from AstraZeneca, Bayer, GSK, Arcus Biosciences, Pyxis, BreakThrough Cancer, Lustgarten Foundation, and NIH/NCI; he has also received honoraria for being on the advisory boards of Partners Therapeutics, BeOne, Prelude, Abdera and Gilead and for serving on the data safety monitoring committee for Astrazeneca and Bayer. **T.K.C.** reports institutional and/or personal, paid and/or unpaid support for research, advisory boards, consultancy, and/or honoraria past 10 years, ongoing or not, from Alkermes, Arcus Bio, AstraZeneca, Aravive, Aveo, Bayer, Bristol Myers-Squibb, Bicycle Therapeutics, Calithera, Caris, Circle Pharma, Deciphera Pharmaceuticals, Eisai, EMD Serono, Exelixis, GlaxoSmithKline, Gilead, HiberCell, IQVA, Infinity, Institut Servier, Ipsen, Jansen, Kanaph, Lilly, Merck, Nikang, Neomorph, Nuscan/PrecedeBio, Novartis, Oncohost, Pfizer, Roche, Sanofi/Aventis, Scholar Rock, Surface Oncology, Takeda, Tempest, Up-To-Date, CME and non-CME events (Mashup Media, Elite Elements, Peerview, OncLive, MJH, CCO and others), Xencor, outside the submitted work. Institutional patents filed on molecular alterations and immunotherapy response/toxicity, rare genitourinary cancers, and ctDNA/liquid biopsies. Equity: Tempest, Pionyr, Osel, Precede Bio, CureResponse, Primium, Abalytics, Faron Pharma. Committees: NCCN, GU Steering Committee, ASCO (BOD 6/2024–), ESMO, ACCRU, KidneyCan. Medical writing and editorial assistance support may have been funded by communications companies in part. No speaker’s bureau. Mentored several non-US citizens on research projects with potential funding (in part) from non-US sources/Foreign Components. The institution (Dana-Farber Cancer Institute) may have received additional independent funding of drug companies or and/or royalties potentially involved in research around the subject matter. **K.J.P.** reports serving on one-time advisory boards for Exelixis, Sanofi, Lilly, and Novartis. **A.J.A.** has consulted for Affini-T Therapeutics, AstraZeneca, Blueprint Medicines, Boehringer Ingelheim, Curie.Bio, Incyte, Kestrel Therapeutics, Merck & Co., Inc., Mirati Therapeutics Inc., Nimbus Therapeutics, Oncorus, Inc., Plexium, Quanta Therapeutics, Reactive Biosciences, Revolution Medicines, Riva Therapeutics, Servier Pharmaceuticals, Syros Pharmaceuticals, Taiho Pharmaceuticals, T-knife Therapeutics, Third Rock Ventures, and Ventus Therapeutics; holds equity in Riva Therapeutics and Kestrel Therapeutics; and has research funding from Amgen, AstraZeneca, Boehringer Ingelheim, Bristol Myers Squibb, Deerfield, Inc., Eli Lilly, Mirati Therapeutics Inc., Novartis, Novo Ventures, Revolution Medicines. **B.M.W.** reports advisory boards/consulting role with Agenus, BeiGene, BMS/Mirati, Cancer Panels, EcoR1 Capital, GRAIL, Harbinger Health, Immuneering, Ipsen, Lustgarten Foundation, Revolution Medicines, SystImmune, Takeda, Tango Therapeutics, Third Rock Ventures; receives research funding to his institution from Amgen, AstraZeneca, Break Through Cancer, Eli Lilly, Harbinger Health, Lustgarten Foundation, Mark Foundation, NIH/NCI, Novartis, Pancreatic Cancer Action Network, Revolution Medicines, Stand Up to Cancer, Stephenson Foundation. **H.S.** reports consulting relationships with Blueprint Medicines, Zola Therapeutics, Dewpoint Therapeutics, and Merck Sharp & Dohme; research support from AstraZeneca; and honoraria from UpToDate. All other authors declare no competing interests.

## Authors’ Contributions

**K.S.:** Conceptualization, formal analysis, software, investigation, methodology, visualization, writing-original draft. **M.E.:** Conceptualization, data curation, investigation, methodology, visualization, writing-original draft. **D.V.:** Resources, data curation, investigation, methodology. **G.G.:** Formal analysis, investigation, methodology. **C.L.:** Resources, investigation, visualization. **E.I.:** Formal analysis, methodology, visualization. **J.-H.S.:** Investigation, project administration. **J.J.C.:** Investigation. **H.Sav.:** Investigation. **A.J.:** Data curation. **L.C.:** Project administration, data curation. **N.P.:** Investigation. **R.N.:** Investigation. **A.S.:** Resources. **A.D.C.:** Investigation. **E.A.A.:** Data curation, project administration. **E.C.C.:** Project administration. **T.E.Z.:** Investigation. **G.G.L.** Methodology. **R.H.C.:** Methodology. **Z.Z.:** Methodology. **G.N.:** Investigation. **W.D.K.:** Investigation. **B.J.** Investigation. **Z.J.** Methodology. **P.C.:** Methodology. **B.F.:** Methodology. **D.P.:** Resources. **C.V.:** Resources. **T.H.:** Resources. **A.H.:** Resources, writing-review and editing. **A.I.:** Resources, writing-review and editing. **B.M.H.:** Resources, writing-review and editing. **J.M.C.:** Resources, Writing-review and editing. **J.E.B.:** Resources, writing-review and editing. **T.K.C.:** Resources, writing-review and editing. **K.P.:** Resources, writing-review and editing. **J.N.:** Resources, writing-review and editing. **A.J.A.:** Resources, writing-review and editing. **B.M.W.:** Resources, writing–review and editing. **S.C.B.:** Resources, formal analysis, methodology, supervision, writing-review and editing. **M.L.F.:** Resources, investigation, methodology, supervision, writing-review and editing. **H.S.:** Conceptualization, formal analysis, resources, supervision, funding acquisition, project administration, writing-review and editing. All authors reviewed and approved the final manuscript.

## Acknowledgements

**G.S.G.** is supported by an NIH T32CA009172 award from the National Cancer Institute (NCI). **J.E.B.** is supported by the US Department of Defense (W81XWH-20-1-0118, HT9425-23-1-0048). **S.C.B.** is supported by the Damon Runyon Cancer Research Foundation, the Fund for Innovation in Cancer Informatics, the Wong Family Award in Translational Oncology, and U01 CA296432 from the National Cancer Institute. **H.S.** is supported by K08CA286749 from the National Cancer Institute.

## Supplementary Materials

**Figure S1.**
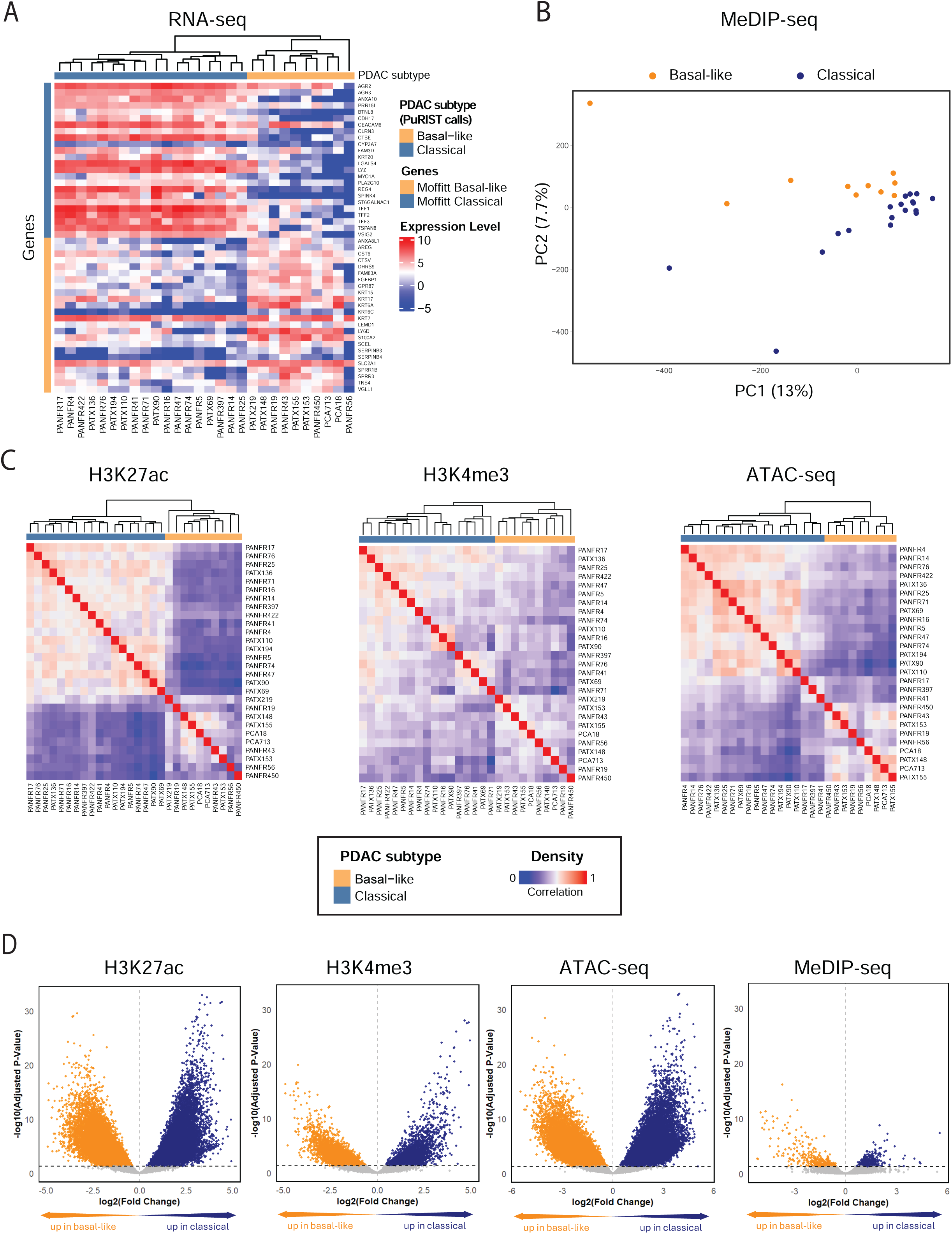
**(A)** Hierarchical clustering of PDAC patient-derived xenograft (PDX) models based on the expression levels of Moffitt subtype–specific genes. PDAC subtypes are indicated by the horizontal color bar and annotated according to PurIST subtype predictions. **(B)** Principal component analysis (PCA) plots of the MeDIP peaks in classical and basal-like PDAC PDX. **(C)** Unsupervised hierarchical clustering of the H3K27ac, H3K4me3, and ATAC peaks. **(D)** Volcano plot of subtype specific differential H3K27ac, H3K4me3, ATAC and MeDIP peaks.

**Figure S2.**
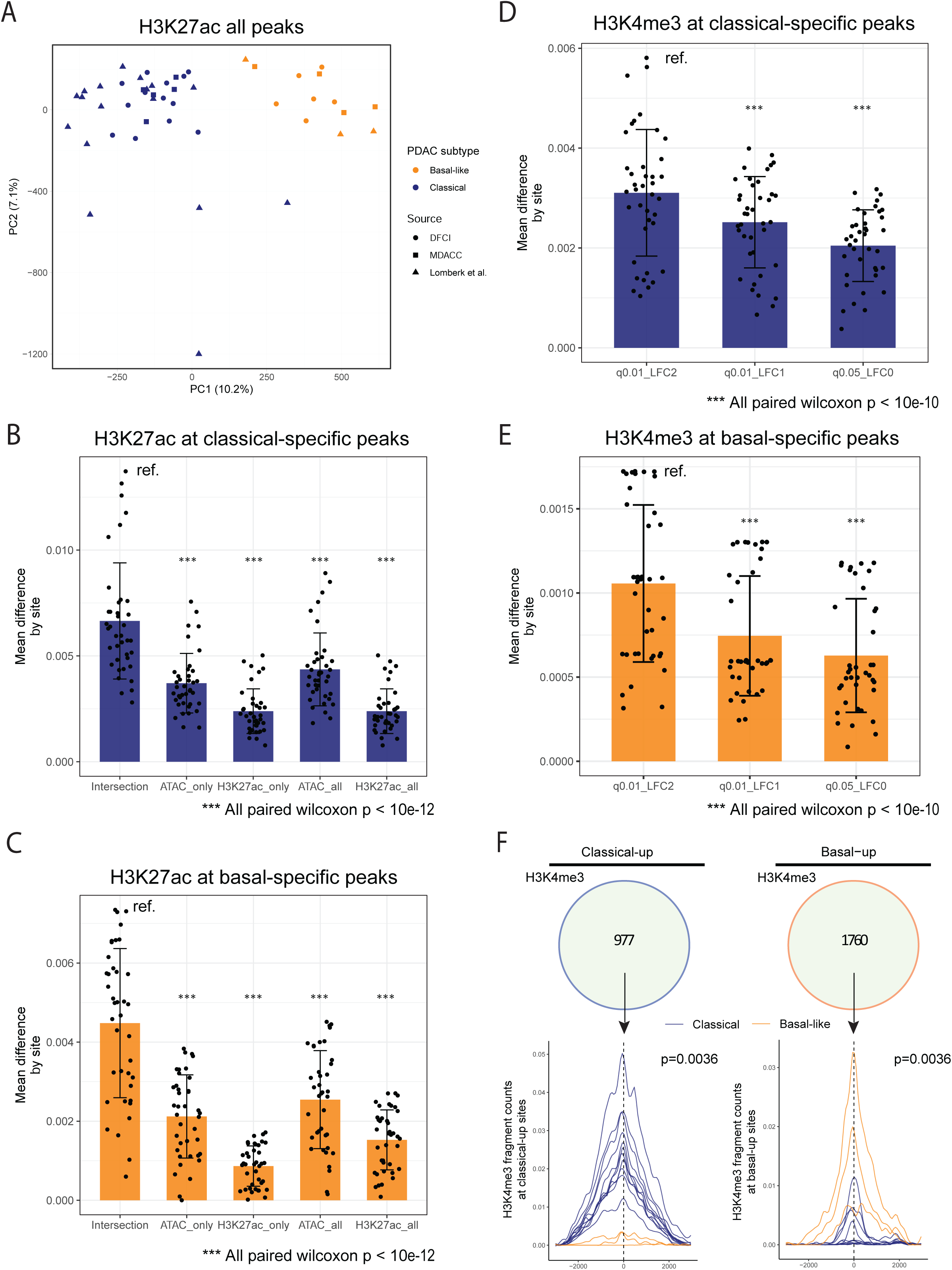
**(A)** Principal component analysis (PCA) of H3K27ac peaks, including the integration of an independent PDX cohort. **(B)** H3K27ac enrichment difference between classical and basal-like PDAC PDXs at classical-specific sites, compared across site sets defined by H3K27ac-only peaks, all H3K27ac peaks, ATAC-only peaks, all ATAC peaks, and the intersection of H3K27ac and ATAC peaks. **(C)** H3K27ac enrichment difference between classical and basal-like PDAC PDXs at basal-specific sites, compared across site sets defined by H3K27ac-only peaks, all H3K27ac peaks, ATAC-only peaks, all ATAC peaks, and the intersection of H3K27ac and ATAC peaks. Intersection: Intersection of H3K27ac and ATAC sites. ATAC only: Regions that are differentially accessible but not differentially marked by H3K27ac. H3K27ac only: Regions that are differentially marked by H3K27ac but not differentially accessible. ATAC all: All differentially accessible regions between classical and basal-like subtypes. H3K27ac all: All differentially marked by H3K27ac regions between classical and basal-like subtypes. **(D)** H3K4me3 enrichment difference between classical and basal-like PDAC PDXs at classical-specific sites, compared across site sets defined by different Log2 fold change (LFC) thresholds. **(E)** H3K4me3 enrichment difference between classical and basal-like PDAC PDXs at basal-specific sites, compared across site sets defined by different LFC thresholds. **(F)** Profile plot showing aggregated H3K4me3 signal across H3K4me3 differential regions at the single-sample level.

**Figure S3.**
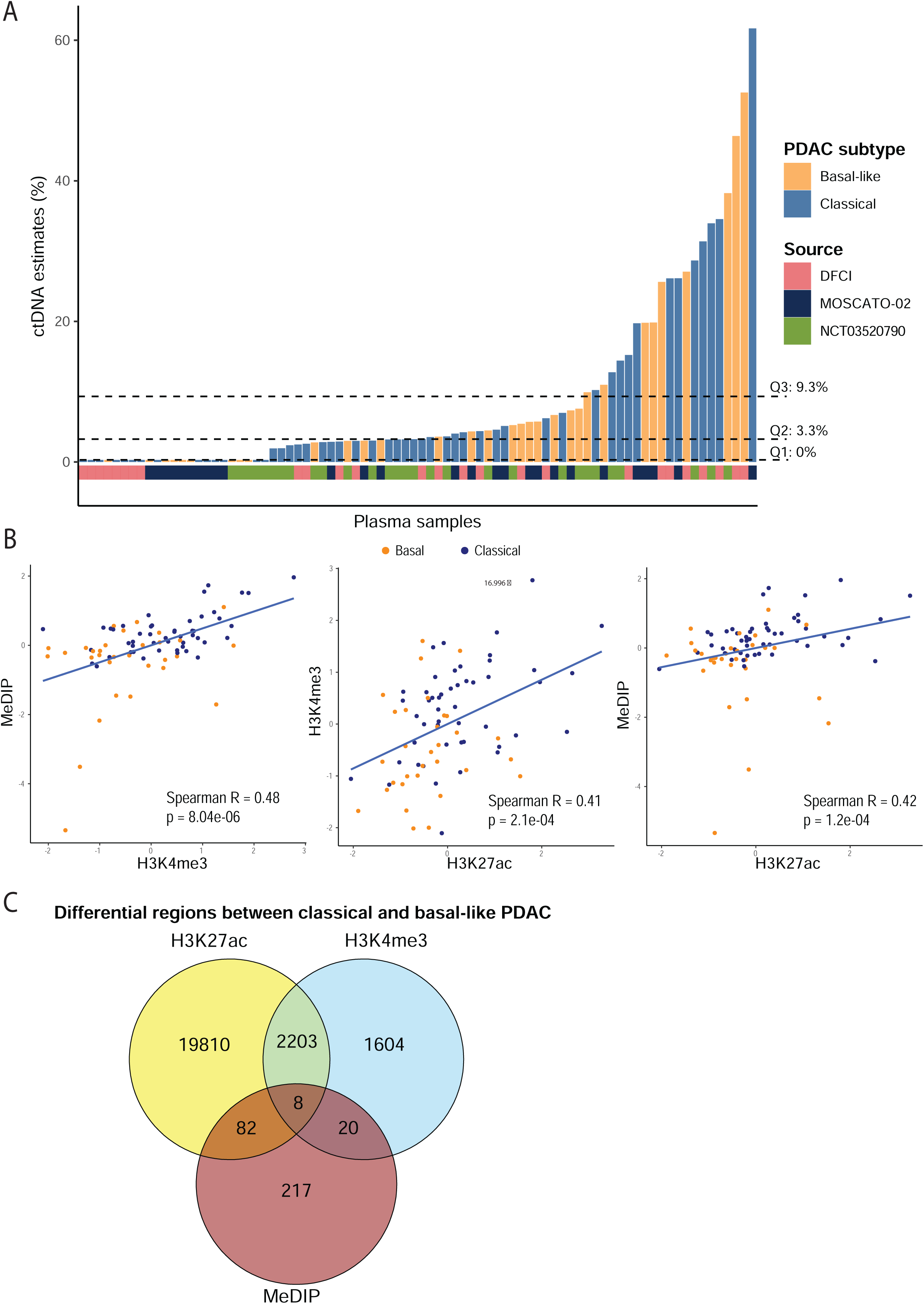
**(A)** Distribution of circulating tumor DNA (ctDNA) fraction, estimated using ichorCNA, in plasma samples from patients with pancreatic ductal adenocarcinoma (PDAC), stratified by transcriptomic subtypes and sample sources. Q1: first quartile; Q2: median; Q3: third quartile. **(B)** Scatter plots of C:B signal ratios derived from cfMeDIP-seq, H3K4me3 cfChIP-seq, and H3K27ac cfChIP-seq in plasma samples from patients with advanced PDAC. Spearman correlation coefficients and corresponding p-values are shown. **(C)** Venn diagram showing the overlap of PDAC molecular subtype-specific differential regions for H3K27ac, H3K4me3, and DNA methylation used for epigenomic ratio calculation.

**Figure S4.**
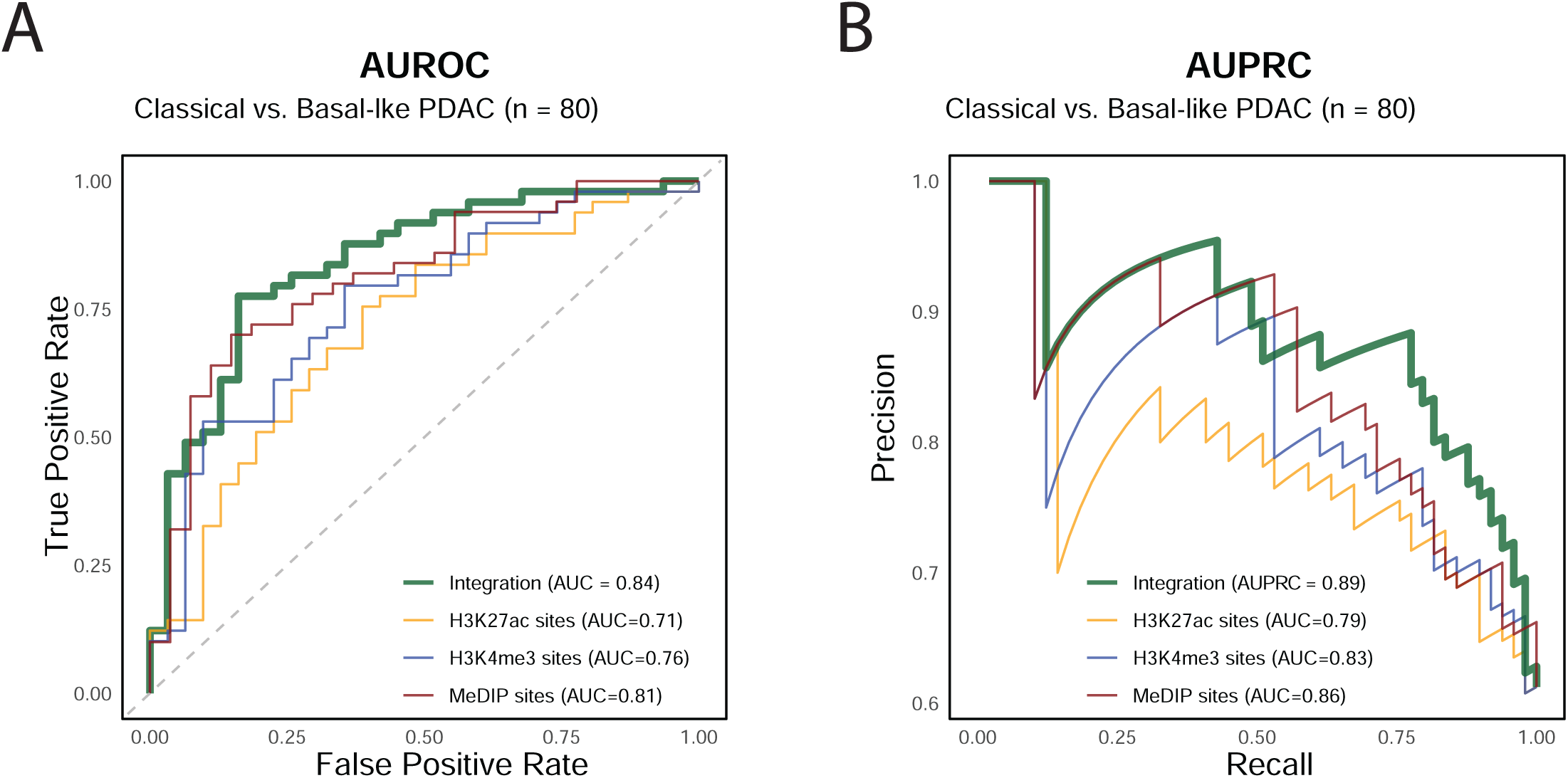
**(A)** Area under the receiver operating characteristic (AUROC) curve for distinguishing classical and basal-like pancreatic cancer by epigenomic mark and their integration. **(B)** Area under the precision-recall curve (AUPRC) for distinguishing classical and basal-like pancreatic cancer by epigenomic mark and their integration.

**Figure S5.**
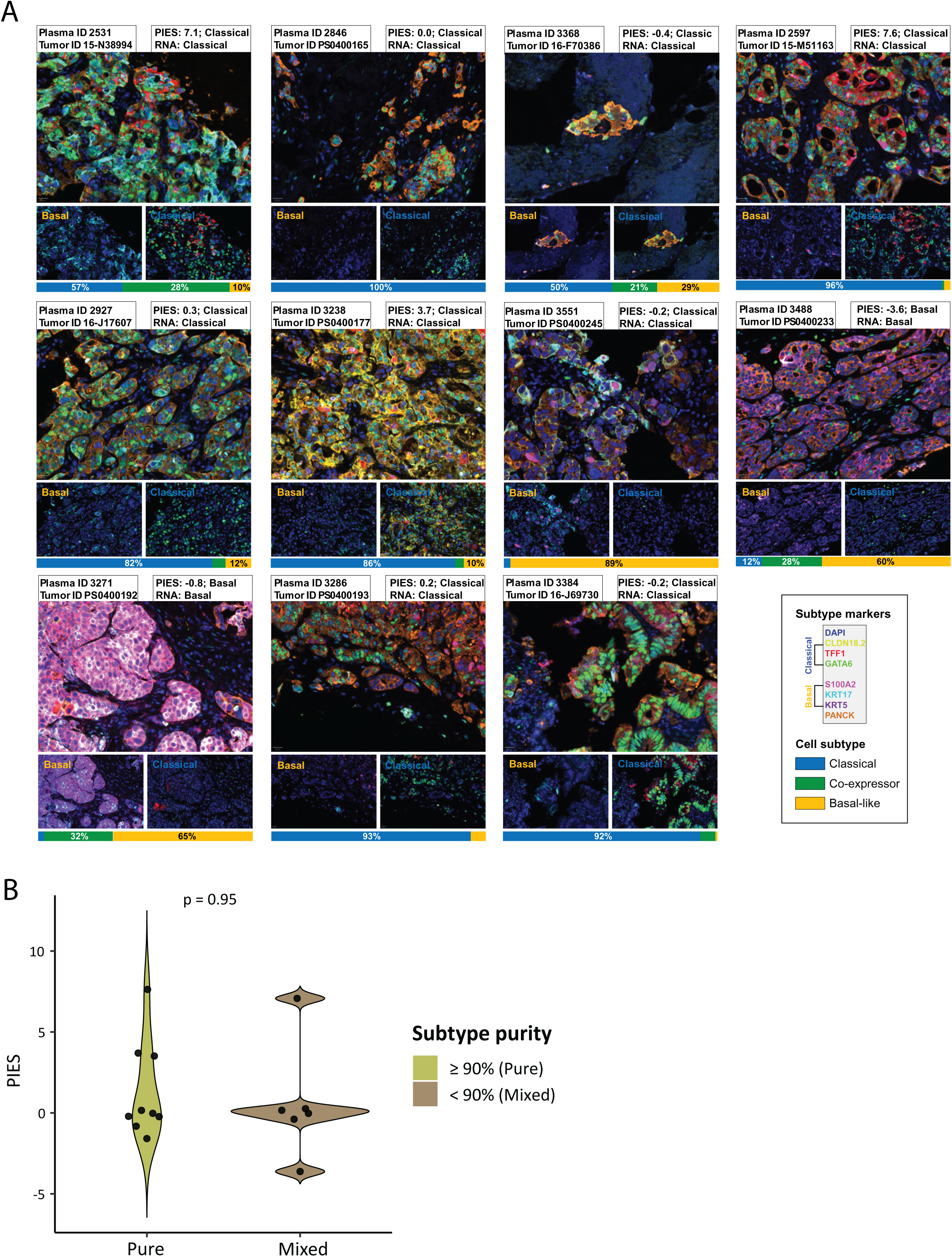
**(A)** Representative mIF images from eleven pancreatic cancer tumors. Scale bar = 20 μm. **(B)** Distribution of PIES scores according to PDAC subtype purity defined by multiplex immunofluorescence (mIF). Tumors classified as mixed (<90% of either classical or basal-like cells by mIF) or pure (≥90% of a single subtype).

**Figure S6.**
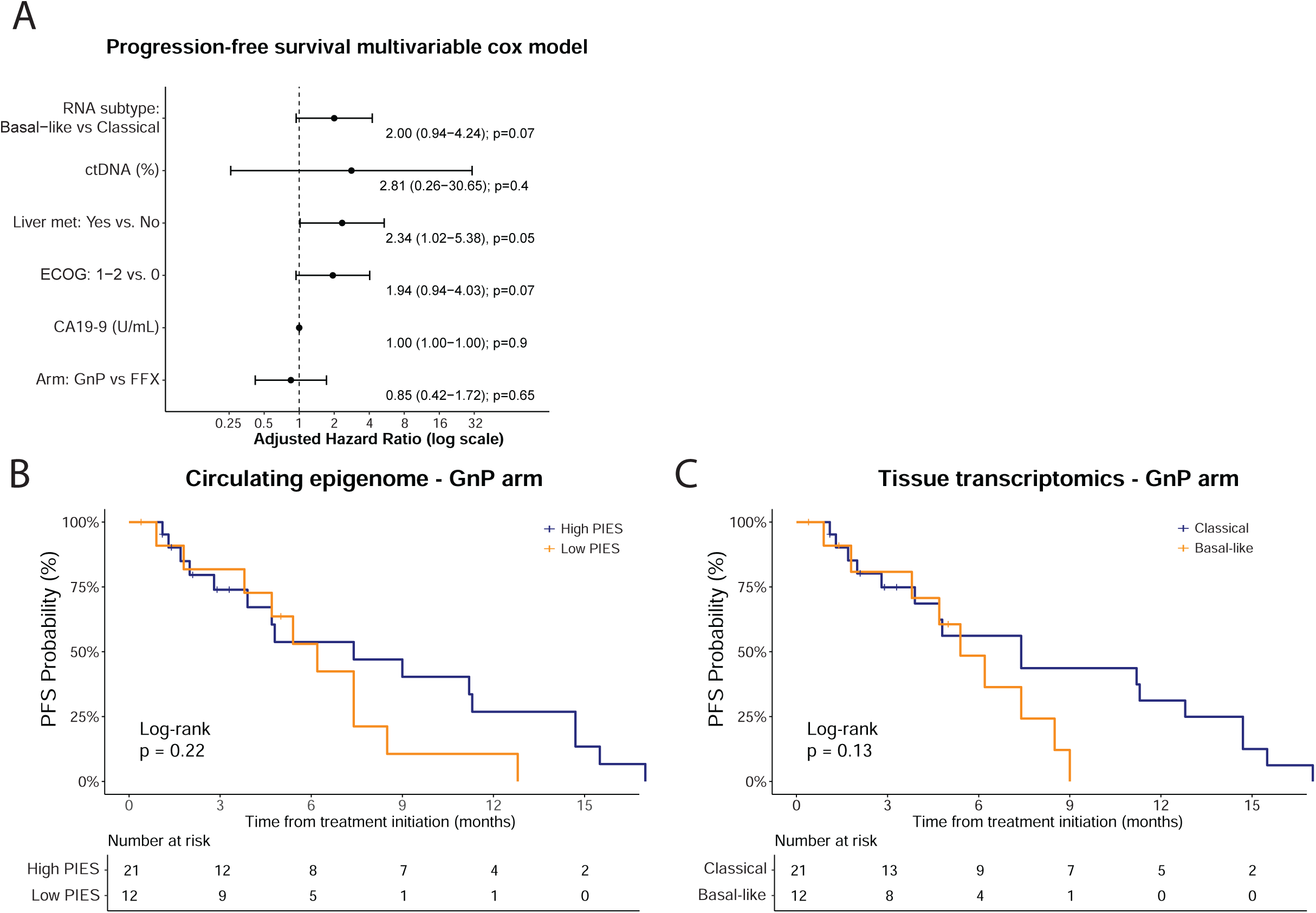
**(A)** Forest plot for multivariable Cox proportional hazards regression model for progression-free survival of patients with metastatic PDAC. **(B)** Kaplan-Meier curves of progression-free survival of patients with metastatic PDAC receiving gemcitabine/nab-paclitaxel (GnP) stratified by high and low PIES. **(C)** Kaplan-Meier curves of progression-free survival of patients with metastatic PDAC receiving GnP stratified by tissue transcriptomics subtypes.

**Supplementary Table 1.** PDAC PDX cohort, quality metrics of generated epigenomic libraries, and subtype calls by PurIST.

**Supplementary Table 2.** List of genomic loci differentially enriched for H3K27ac, H3K4me3, ATAC, and MeDIP in classical and basal-like PDAC.

**Supplementary Table 3.** List of external PDAC PDX dataset with subtype calls by PurIST.

**Supplementary Table 4.** Demographics of patients in the metastatic PDAC plasma cohort and quality metrics of generated epigenomic libraries.

**Supplementary Table 5.** Sample-level comparison of PDAC subtyping across liquid biopsy (PIES) and tissue-based (PurIST and multiplex immunofluorescence (mIF) subtypes) platforms.

## Notes

### Clinical Trial

NCT03520790 and NCT01566019

### Author Declarations

IRB of Dana-Farber Cancer Institute gave ethical approval of this work. All procedures were conducted in accordance with institutional guidelines, HIPAA regulations, and the Declaration of Helsinki. Additional approval was obtained from participating institutions as required.

